# Multi-omic spatial effects on high-resolution AI-derived retinal thickness

**DOI:** 10.1101/2023.07.31.23293176

**Authors:** VE Jackson, Y Wu, R Bonelli, J Owen, S Farashi, Y Kihara, ML Gantner, C Egan, KM Williams, BRE Ansell, A Tufail, AY Lee, M Bahlo

**Author notes:** Corresponding author: Melanie Bahlo. = joint first authors (VE Jackson, Y Wu, R Bonelli). = joint last authors (AY Lee, M Bahlo).

## Abstract

Retinal thickness is a marker of retinal health and more broadly, is seen as a promising biomarker for many systemic diseases. Retinal thickness measurements are procured from optical coherence tomography (OCT) as part of routine clinical eyecare. We processed the UK Biobank OCT images using a convolutional neural network to produce fine-scale retinal thickness measurements across >29,000 points in the macula, the part of the retina responsible for human central vision. The macula is disproportionately affected by high disease burden retinal disorders such as age-related macular degeneration and diabetic retinopathy, which both involve metabolic dysregulation. Analysis of common genomic variants, metabolomic, blood and immune biomarkers, ICD10 codes and polygenic risk scores across a fine-scale macular thickness grid, reveals multiple novel genetic loci-including four on the X chromosome; retinal thinning associated with many systemic disorders including multiple sclerosis; and multiple associations to correlated metabolites that cluster spatially in the retina. We highlight parafoveal thickness to be particularly susceptible to systemic insults. These results demonstrate the gains in discovery power and resolution achievable with AI-leveraged analysis. Results are accessible using a bespoke web interface that gives full control to pursue findings.

**Graphical Abstract:** 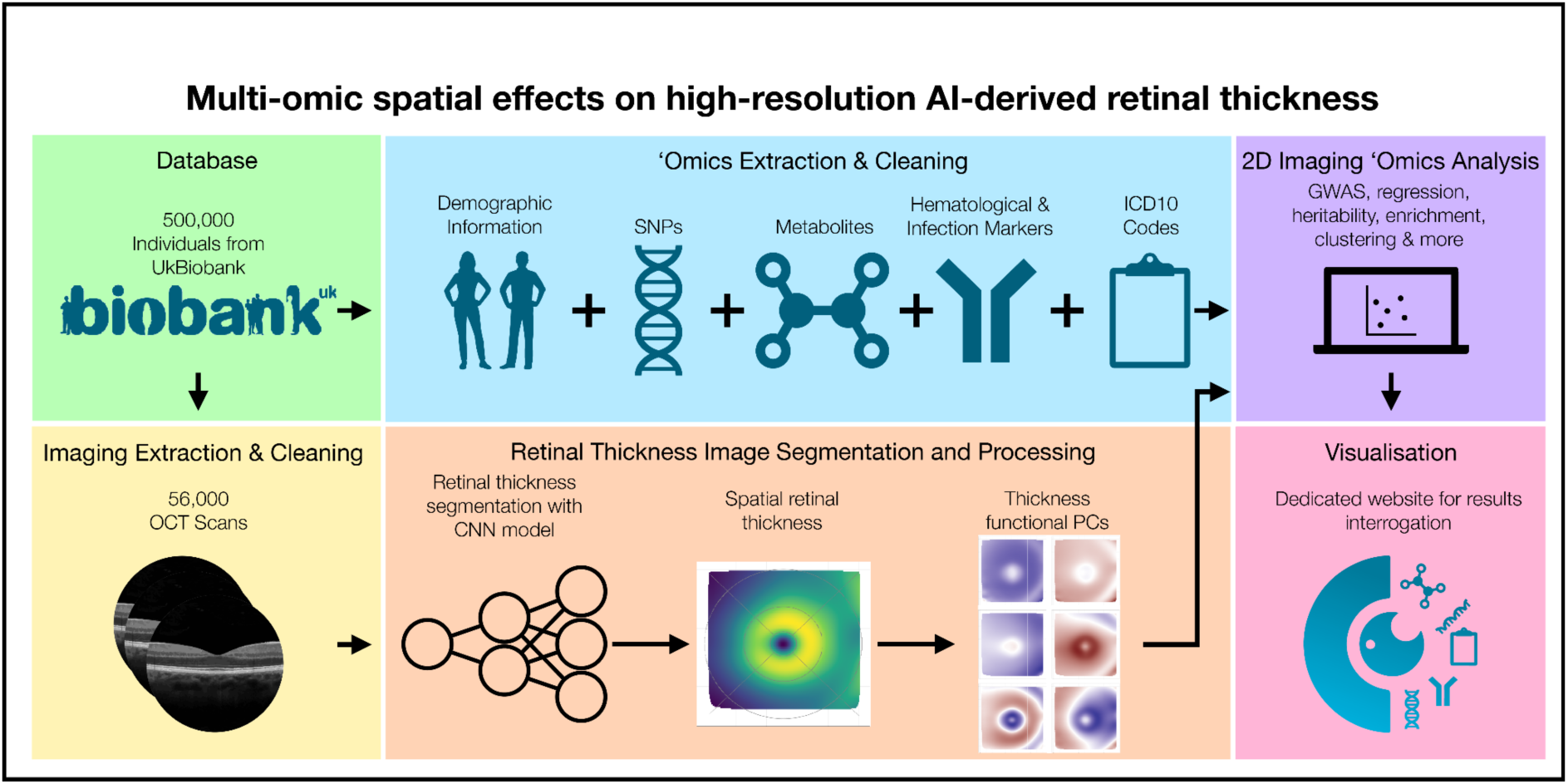

## Main

The retina comprises multiple anatomical layers of more than ten types of highly specialised cells that enable sight ^1, 2^, and features clinically relevant zones such as the macula, fovea and optic nerve. Many hereditary and acquired human retinal conditions show a predilection for the macula, such as age-related macular degeneration (AMD), diabetic retinopathy and macular telangiectasia Type 2 (MacTel).

Optical Coherence Tomography (OCT), is a widely used imaging method that extracts information about retinal morphology, including overall retinal thickness (RT). Retinal thickness is a composite measure across several sublayers, and is influenced by multiple determinants at any particular location in the macula, such as: amount of vascularisation, atrophy, ischemic damage, and presence or absence of particular cell types, such as rod and cone cells. RT is highest in the perifoveal region, dipping to a minimum at the foveal pit, located at the center of the macula, an area enriched for cone cells, yet depleted for rod cells. Such substantial morphological variation over such a small area suggests the presence of tightly spatially regulated biological processes.

The retina is part of the central nervous system, empirically confirmed with transcriptomics clustering of retina with brain^3^. The field of ‘oculomics’ is growing rapidly, with AI-enabled retinal imaging based disease prediction models being developed for neuropsychiatric disorders such as schizophrenia^4^ and for non-neurological disease such as cardiovascular disease^5^, with promise for preventative health care. RT also has diagnostic potential for many diseases, particularly neurodegenerative disorders such as dementia and Parkinson’s disease^6^ and multiple sclerosis, where it was first proposed as a potential biomarker in 2011^7^.

Previous genome-wide association studies (GWAS) of retinal OCT data from UK Biobank (UKBB) made use of the imaging platform’s associated algorithm (TOPCON/TABS), with measures, summarized over the widely used Early Treatment of Diabetic Retinopathy Study (ETDRS) or macula 6 grids, for overall RT ^8^ and retinal sublayers ^9, 10^. Here, we processed retinal OCT data from UKBB, with a deep learning based image segmentation method to produce a high resolution RT dataset, which we used to investigate the relationships between RT and genetic variation, in addition to metabolites, blood traits, immunological traits and disease (Figure 1).

**Figure 1.**
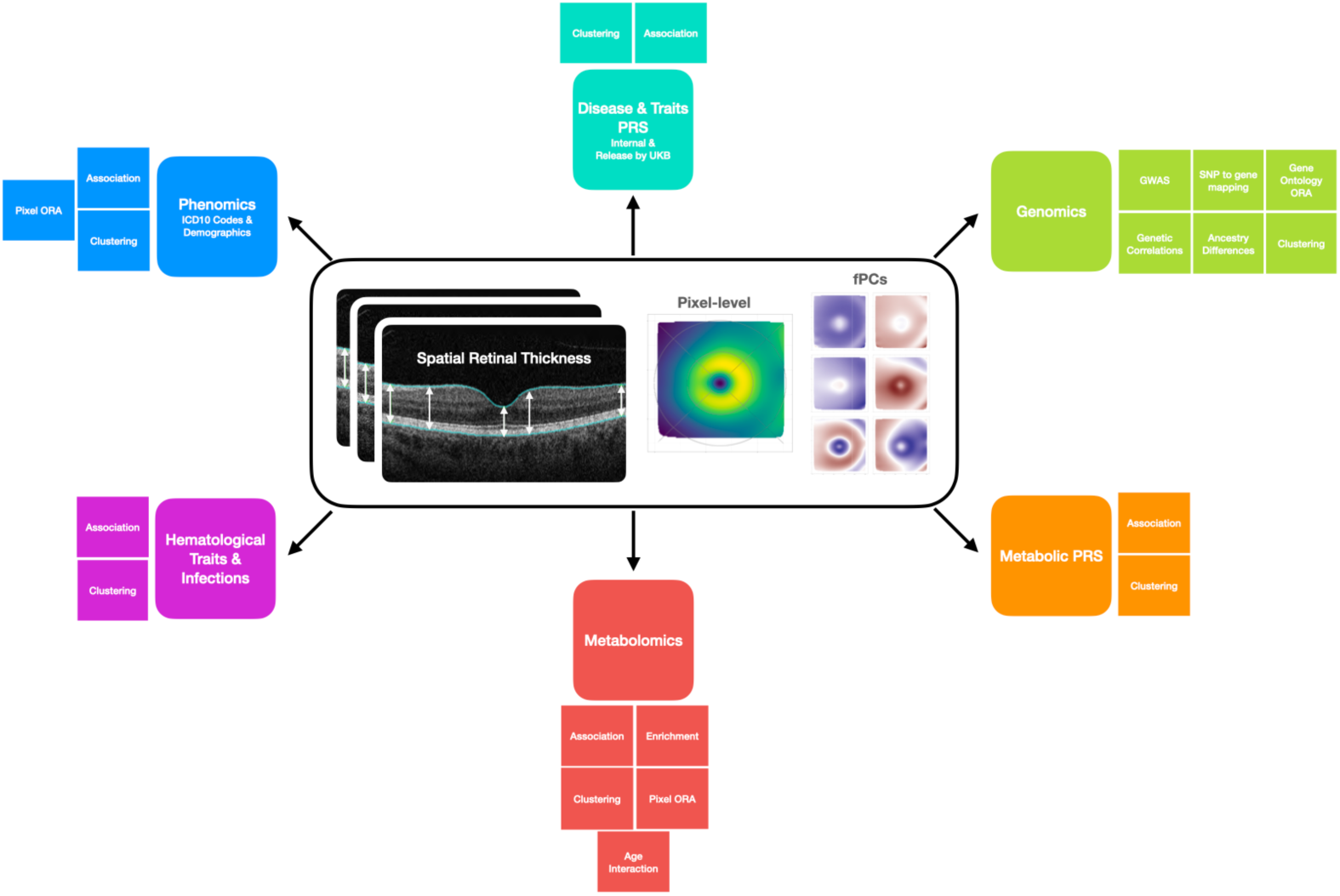
Retinal thickness definition and analyses. Definition of retinal thickness used as the primary outcome measure, based on retinal thickness produced by a CNN. RT captures the thickness between the internal limiting membrane and the retinal pigment epithelium with schema of analyses performed.

Results reveal why retinal imaging is ‘a window to the brain’, by detecting the connections to neurological and vascular disorders, and pinpointing specific regions of the macula driving these associations. We highlight the complex spatial anatomy of the macula, providing context for novel candidate genes and their biological mechanisms. Our rich multi-omics results are easily accessible in their entirety through a bespoke interactive browser (https://retinomics.org/), and set a new benchmark for complex, two dimensional image analyses in population samples.

## Results

### Retinal Thickness Imaging Data

Retinal thickness is defined as the distance between, as well as including, the inner limiting membrane (ILM) and retinal pigment epithelium (RPE) layers (Figure 1, inset).

OCT imaging data for at least one eye was available from 85,793 UKBB participants. All individuals with evidence of retinal diseases were excluded from analyses (Supplementary Table 1) but individuals with other non-retinal eye diseases or self-reported vision problems, in the absence of retinal disease, were retained.

OCT images were filtered for overall quality and then processed using a deep convolutional neural network (DCNN), based on a method used in ^11^, to produce RT estimates over a grid of 128 (superior/inferior axis) by 256 (temporal/nasal axis) pixels, followed by further QC (Supplementary Figures 1-6). Each pixel captured approximately 46.88 by 11.72 μm^2^ of the retina and the total area analyzed captured 6000 by 6000 μm^2^, with the fovea at the center. Images from the left side were reflected around the foveal midpoint to be anatomically aligned to the right side for further analysis (Supplementary Figure 7). RT measurements are expressed as pixels, where a pixel corresponds to approximately 3.5μm.

Extensive quality control of the RT data removed individuals, scans, and pixels with poor data (Supplementary Figure 8). Missing pixel values were imputed using a generalized additive model to produce a complete dataset for RT for all individuals, over 29,041 pixels. Individuals with data available from both eyes had their RT values averaged such that each individual only had a single set of RT values for all subsequent analyses. The final dataset included 54,844 participants with OCT data, with 36,653 with images for both eyes and 18,191 with images for one eye only.

### Functional Principal Component analysis captures broad spatial patterns

Functional principal component (fPC) analysis is a dimensionality reduction method, akin to principal component analysis, which estimates the primary modes of variation of functional data, such as curves or surfaces. We applied this approach using tensor product splines as base functions, using the MFPCA R package.^12, 13^ Scree plots revealed that six fPCs captured ∼95% of the RT variation (Supplementary Figure 9), each capturing striking spatial patterns: fPCs 1 and 2 capture variation in the perifovea, and outer retinal regions; fPC3 corresponds to thickening on the nasal side; fPC4 represents reduced parafoveal thickness; fPC5 captures the shape of the foveal pit; and fPC6 denotes thickening of the parafovea and nasal side (Extended Data 1). For each individual, fPC scores were extracted, representing the contribution of each fPC to that individual’s scan.

### Overview of analysis cohort

For all analyses, the cohort was restricted to unrelated individuals, whose broad continental ancestries were European (EUR, n=43,148 individuals), Central and South Asian (CSA, n=1,179) or African (AFR, n=1,161), based on genetic similarity to individuals from the 1000 Genomes Project and Human Genome Diversity Panel. The outcome RT datasets used for analyses consisted of 29,041 pixels (pixel-level analyses) and six fPC scores (fPC analyses), defined above. Summary RT measures, and basic characteristics for the analysed cohort are given in Supplementary Table 2 and Extended Data 2, stratified by ancestry.

We examined RT (fPCs and pixel-level) for relationships with characteristics previously shown to be associated with RT. On the pixel-level, we recapitulated previously reported effects for age, sex and spherical equivalent (Supplementary Figure 10) ^14^. Males had higher RT values than females, particularly in the fovea and parafovea. Age had a non-linear relationship with RT, with a maximum at 54 years, then thinning with age. Higher values of spherical equivalent (greater hyperopia) showed highly significant associations with thicker RT values in the para- and perifovea, and more modest associations with thinner RT values within the fovea. In addition, increased standing height, as a proxy for body size, was associated with thicker RT in the parafovea. FPCs were also associated with these factors, to differing degrees (Supplementary Table 3).

For the subsequent multi-omics analyses, all associations with RT were investigated with adjustment for age, age-squared, sex, standing height, spherical equivalent and ancestry (10 ancestry Principal Components), with device and eye (left, right, both) as technical covariates. The multi-omics study data analysis plan is summarized in Figure 1, with sample sizes for each analysis in Supplementary Table 4.

### Genetic association analyses

GWAS were undertaken using genotypes imputed to the combined Haplotype Reference Consortium and UK10K panel ^15^. The primary discovery analyses were conducted on the EUR ancestry individuals, with secondary analyses on the CSA and AFR individuals, to assess ancestral heterogeneity. Associations with 11,239,006 SNPs from the 22 autosomes and the X chromosome, were examined using PLINK 2 (version 20221024). Consolidation of all GWAS results from across the 29,041 pixel-level and 6 fPCs was achieved using a post-hoc iterative procedure that incorporated genomic spatial correlation, via LD clumping, to allocate SNPs and RT pixels/fPCs to independent loci (Supplementary Figure 11). We report loci meeting Bonferroni corrected genome-wide significance thresholds for each approach: P<5E-8 / 29,021 = 1.72E-12 for the pixel-level analyses; P <5E-8 / 6 = 8.33E-9 for the FPC analyses.

### Number and distribution of loci identified through both fPC and pixel-level analysis

We identify 224 unique RT-associated genetic loci that meet the pixel-level Bonferroni corrected threshold (Supplementary Table 5). The number of loci that achieve significance for each pixel has a symmetric distribution with a mean of 22.8 loci (sd = 5.7, min = 4, max = 42) per pixel (Extended Data 3). The density of the number of loci forms concentric rings mainly concentrated in the foveal region, suggesting changing patterns of association. These rings may represent the echoes of retinal development ^16^. The fPC-based RT association analyses identified 120 loci (Supplementary Table 6), with 4, 11, 36, 45, 42, 23 loci meeting P < 8.33E-9 for FPCs 1 to 6, respectively (Extended Data 4). A total of 294 RT genetic loci were identified collectively through either the pixel-level and fPC analyses, with 92 identified through both approaches. All loci were mapped to candidate genes via positional mapping, eQTL data, and chromatin interaction data (Supplementary Tables 7-10). Associations and candidate genes for each of the RT loci are depicted in Figure 2.

**Figure 2.**
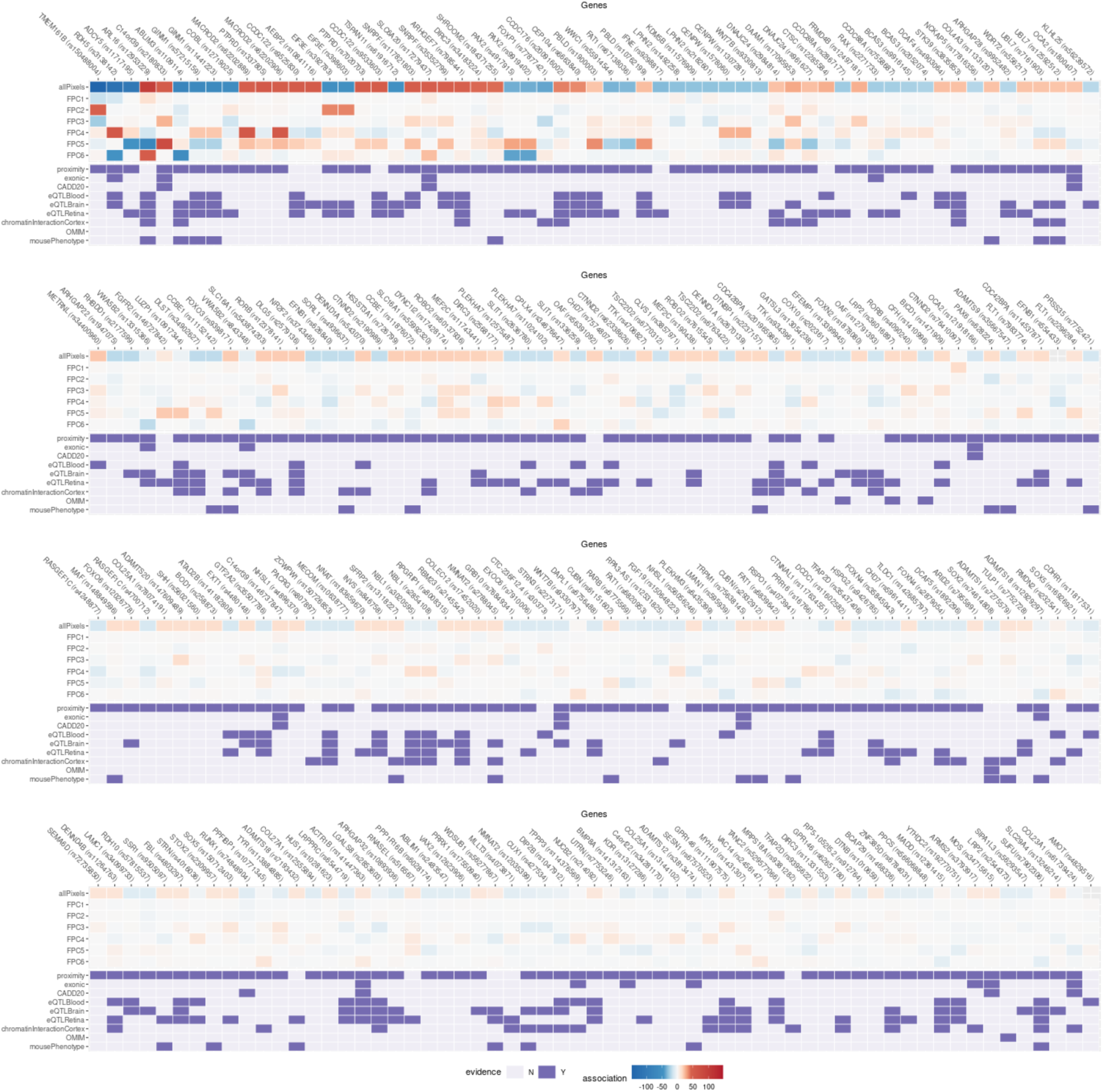
Prioritized genes from GWAS. Summary results for loci identified through the pixel-level and fPC GWAS. For each locus, the upper seven rows summarize the strength and direction of the genetic association based on the beta value direction and the -log10 p-value (Bonferroni adjusted p-values shown to allow comparison across pixel-level and fPC results). The bottom 9 rows summarize locus to gene mapping evidence, based on proximity (<5kB), presence of variants that are exonic; with CADD score > 20; eQTLs; or in chromatin-interacting regions. Implicated genes with retina-associated phenotypes in OMIM and/or in mouse models, are also indicated.

A majority of the loci identified in the Gao et al 2018 ^8^ analyses reached the Bonferroni corrected significance threshold in the pixel-level and/or fPC analyses (89/140, 64%), with an additional 31 loci meeting P<5E-8 (Supplementary Figure 12). The concordance with the Currant et al. 2021 ^10^ and Currant et al. 2023 ^9^ loci was lower with 25/47 and 52/111 identified through our approach, respectively.

The top two loci from our pixel-level analyses were amongst those identified previously: rs150408004 (pixel-wise minimum p = 3.94e-152, beta = 2.03; fPC2 p=6.54e-46), is a long distance enhancer SNP affecting *LINC00461*, a long non-coding RNA now known to be integral for early Müller glial cell and astrocyte development in the human retina ^2, 17^ and which has been previously identified in multiple macula phenotype GWAS studies ^18, 19^; and rs3138142, coding for a synonymous SNP in *RDH5* (pixel-wise minimum p = 3.27e-125, beta = 0.82; fPC4 = 1.06e-58), which codes for the visual cycle enzyme 11-cis retinol dehydrogenase 5, essential for night vision, with mutations in this gene causing a retinal phenotype called fundus albipunctatus (OMIM #136880).^20^ Clustering analysis of the sentinel SNP effects from the pixel-level analysis, revealed 10 clusters impacting retinal thickness in a similar manner (Supplementary Figure 13). Clustering of the pixels highlighted concentric regions of the macula similarly affected by genetic perturbations (Supplementary Figure 14).

### Novel RT associations

There was a significant gain in the number of loci beyond previously published work (Extended Data 3), despite the substantial and strict Bonferroni testing correction. We identified 140 novel RT loci, 70 uniquely from pixel-level analysis, 28 uniquely from the fPC analysis, with 42 found in both. Many of these have been previously implicated in GWAS for ocular traits, such as rs77877421 (*FOXP1*, pixel-level minimum p = 8.42e-44, fPC3 p=4.21e-20), previously identified as a GWAS hit for intraocular pressure ^21^ and vertical cup-disc ratio ^22^, both using UKBB. *Foxp1* regulates the timing of early retinal cell production ^23^. Similarly, the *MACROD2* locus (pixel-level sentinel rs62202906, minimum p = 1.93e-54, fPC4 sentinel rs62202889, p=5.78E-58) has been previously identified as a suggestive finding for thyroid-associated orbitopathy, an immune-mediated eye disorder. Interestingly our spatial analysis reveals that the signal mainly originates from the nasal perifoveal region, with thinning effects on the superior quadrant and thickening effects on the inferior quadrant (Figure 5A). rs61916712 (pixel-level minimum p=5.59e-56) showed a strikingly different association pattern, highly focused within the fovea (Figure 5B); this locus overlaps a retinal eQTL for *TSPAN11*, and a GWAS hit for optic disc morphology. ^22, 24^ Another novel signal rs74614808 (pixel-wise min P=5.60E-15) was detected in a region with a significant chromatin interaction with *SOX2* in adult and fetal cortex. Variants in *SOX2* cause microphthalmia, a birth defect in which one or both eyes fail to fully develop, with optic nerve hypoplasia and abnormalities of the central nervous system.^25^ *Sox2* KO mice have abnormal optic disc and retina blood vessel morphology. ^26^

Amongst the novel findings were the first four X chromosome loci found to be associated with RT. Two of these have clear retinal roles: rs626840 (*EFNB1*, fPC4 p=3.28e-15), and rs554433 (pixel-level p=1.82e-24), an intronic variant in *SHROOM2*, a gene involved in pigmentation of the retinal pigment epithelium, and rs626840 in *EFNB1*. Mutations in *EFNB1* cause craniofrontonasal syndrome, an X-linked developmental malformation, which is reported to cause a number of ophthalmologic abnormalities, including strabismus, nystagmus and hypermetropia ^27^ and has been linked to glaucomatous optic neuropathy ^28^. The spatial signal indicates the strongest effect in the perifoveal superior/nasal quadrant, the closest region to the optic nerve, with the risk allele leading to retinal thickening.

### Cross-ancestry comparisons, and biological insight

We assessed the effects of all loci identified in our EUR discovery analyses in the AFR and CSA ancestry individuals (Extended Data 5, Supplementary Table 11). In the pixel-level, and fPC analyses, the EUR and CSA effect estimates for the top pixel, or top fPC showed strong correlations (pixel-wise rho=0.618, p= 1.141e-24; fPC rho = 0.630, p = 2.00E-14), with weaker correlations for the EUR and AFR effects (pixel-wise rho = 0.305, p=4.54e-06; fPC rho = 0.249, p = 6.8e-03). For the pixel-level loci, we also compared whether the pattern of association across the entire scan was similar across ancestries. The median scan-wise correlation (i.e., the correlation of effects across all pixels), for EUR vs CSA individuals was rho = 0.306, with 28.0% of sentinel SNPs having rho >= 0.5. Again, concordance was lower in EUR vs AFR (Median correlation: rho = 0.195; 11.5% sentinels with rho >= 0.5).

Of the loci identified through our discovery pixel-level analysis, four were significantly associated (P < 0.05 / 224) with RT in CSA individuals: rs3138142 (*RDH5*); rs10164933 (*LINC01248*); rs183659670 (*NNAT*); rs1109114 (*ABLIM3*), with the latter two being novel loci. Of note, rs183659670 is associated with *NNAT* expression in the retina (r^2^=1 with the top eQTL SNP); this gene encodes a proteolipid involved with brain and nervous system development. In the fPC analysis, 2 SNPs met the Bonferroni corrected threshold (P < 0.05 / 120) for CSA individuals, both are most significantly associated with fPC5. rs1900003 near *PBLD* and *ATOH*; the latter gene encodes a member of the basic helix-loop-helix family of transcription factors, and is thought to play a role in retinal ganglion cell and optic nerve formation, with mutations in this gene causing nonsyndromic congenital retinal nonattachment.^29^

We examined genetic correlations between our fPC genome-wide results, and a range of traits and diseases (Extended Data 6, Supplementary Table 12). The most statistically significant correlations were between fPC5 and urate levels, heart failure, reticulocytes, venous thromboembolism (VTE) and diabetes, and between fPC4 and receipt of disability allowance, a proxy for disability. The traits showing the largest magnitude correlations included ranitidine use (used to treat gastro-oesophageal reflux, fPCs 1 and 5), alcohol-related mental health problems (fPC6), cataract (fPC2), and having received psychiatric care (fPC4). Gene set over-representation analysis was performed mapped genes, implicated via the pixel-level and fPC analyses. Both analyses were enriched for genes involved in eye and visual system development, cell differentiation and kidney development (Supplementary Table 12, Supplementary Figure 15).

### Metabolomic association analyses

325 metabolic measures, from 10,668 participants were included in the association analyses. We examined associations at the single metabolite level and performed hierarchical clustering of both metabolites and pixels to investigate potential spatial effects (Methods). After correction for multiple testing, all metabolites were significantly associated with RT at least one pixel (Supplementary Table 14, Extended Data 7A). A visual representation for the subset of non-derived metabolites is presented in Figure 3. Metabolites calculated as derived lipoprotein ratios were the most significantly associated class of metabolites. The ratio of linoleic acid to total fatty acids (“Linoleic Acid to Total Fatty Acids percentage”) had the strongest positive effect on retinal thickness, while “Phospholipids to Total Lipids in HDL percentage” had the strongest global negative effect.

**Figure 3.**
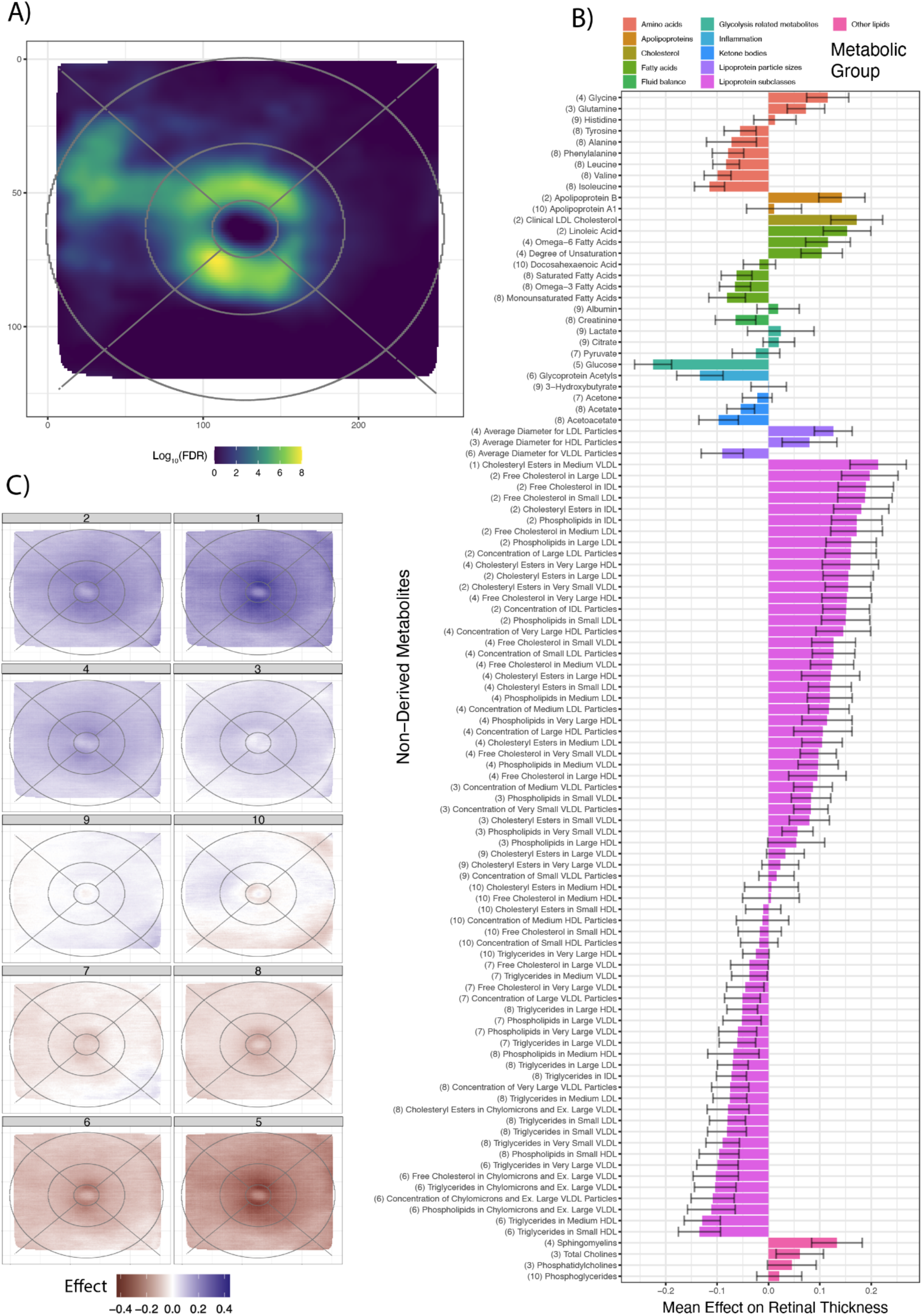
Clustering based on metabolic results. A) Pixel-wise metabolic overrepresentation analysis results B) Average (bar length) and standard devitation (error bars) effect on RT across pixels for the subset of non-derived metabolite. Color represent metabolic group of each metabolite. Cluster number are shown in brackets before each metabolite name. C) Pixel-wise average effect on RT across all metabolites within each detected cluster.

We identified 10 metabolic clusters, each associated with pixels co-located in specific retinal regions (Figure 3, Supplementary Table 14). Clusters 1 and 2 have the highest association with retinal thickness, and contain highly related metabolites, with enrichment for cholesterols (particularly in LDLs (Figure 5D) and VLDLs), apoB, as well as omega 6 fatty acids. Clusters 5, 6 and 8 showed a negative association with RT and include triglycerides, glucose (Figure 5C), branched chain amino acids (BCAA), alanine and chylomicrons.

Multiple metabolites associated with RT are linked to retinal disorders with alterations in systemic metabolites (AMD, MacTel and diabetes). Metabolite changes that are associated with increased AMD risk (lower levels of Cholesterol, LDL, VLDL and apoB, higher levels of triglycerides, BCAA and alanine) ^30, 31^, were found to be associated with retinal thinning. Similarly, changes in glycine, sphingomyelin and alanine that are associated with increased risk of the retinal degenerative disorder, MacTel, were also found to be associated with retinal thinning.^18, 32^ Many of the metabolites negatively associated with retinal thickness; triglycerides, glucose, BCAA and alanine, are elevated in metabolic syndromes and diabetes ^33^, which also correlates with retinal thinning through the often combined effects of retinal neuropathy, changes in vasculature and the occurrence of edema. Unexpectedly, DHA and omega 3 fatty acids, which are important for retinal function, are negatively associated with retinal thickness.^31^

Specific retinal regions were identified as being consistently affected by metabolic disturbances (Figure 3). These matched, to some extent, with recognised retinal anatomical landmarks, as described with the ETDRS grid (Supplementary Figure 7), but also revealed clusters spanning multiple sectors of the grid. Pixel overrepresentation analysis (ORA) revealed that the parafovea, particularly its lower temporal side, was susceptible to metabolic influences (Figure 3C). This was confirmed by the many metabolites significantly affecting fPC4 (Extended Data 7B), which captured parafoveal thickness variation. The temporal perifoveal region was also specifically impacted by metabolic dysregulation.

Age - metabolite interaction analyses, revealed a generalized increase in metabolic effect on retinal thickness with age (Supplementary Table 15). ORA revealed that pixels located to the parafoveal area were again the strongest contributors to the interaction effects on RT (Extended Data 7C).

Metabolic polygenic risk scores (PRSs) have recently been endorsed as a tool for trait associations.^34^ Associations with PRS for Phosphatidylcholines and their lyso-phosphatidylcholines were associated with retinal thickness, as well as certain amino acids such as lysine, glycine and threonine (Supplementary Table 16, Extended Data 8D). All three of these amino acids mainly affected the parafoveal areas.

### ICD10 disease code and comorbidity class analysis

Association with 1,496 ICD10 diagnoses were examined in 36,196 participants. 459 diseases (30%) were significantly associated with RT (Supplementary Table 17, Figure 4B), with the majority (80%) presented as a thinning effect as well as effects on multiple fPCs (Figure 4C). More generally, even among those non-significant, there was a negative relationship between disease status and retinal thickness.

**Figure 4.**
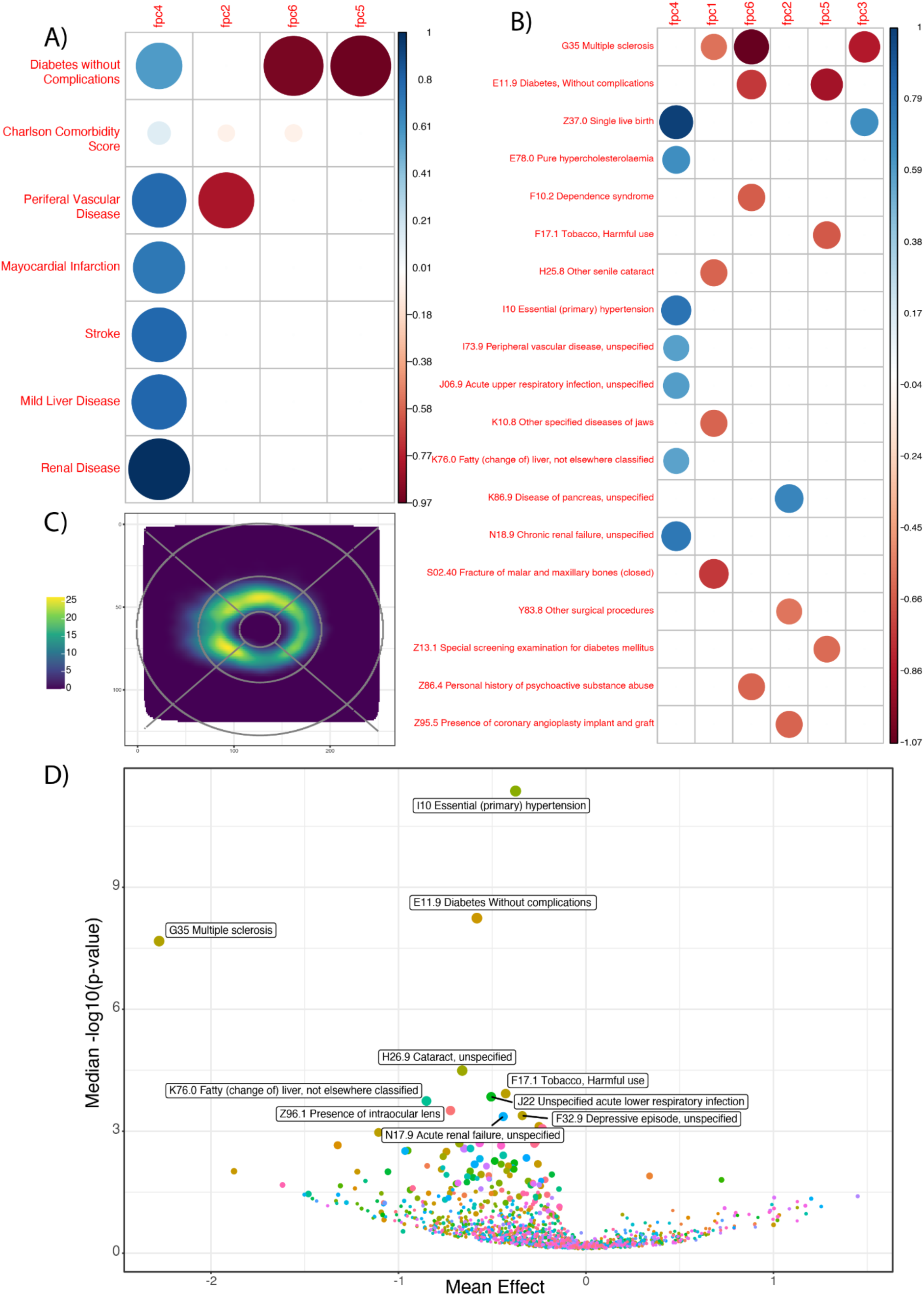
ICD10 diagnoses and comorbidity results. A) Heatmap showing significant association between comorbidity scores and thickness fPCs. B) Heatmap showing significant association between ICD10 diagnoses and thickness fPCs. C) 2D smoothed results from ORA analysis on main ICD10 results. D) volcano plot showing average effect size across retinal thickness and median -log10(p-value) for each ICD10 diagnosis.

Multiple sclerosis (MS, G35, N = 133 cases reported in UKBB, mean effect size for RT for all significant log adjusted p-value values across all pixels = -2.46) had the largest significant global effect, with strongest effects observed in the nasal perifoveal region closest to the optic disc (Figure 5 E). MS results in oligodendrocyte demyelination, with impacts on the optic nerve as previously reported.^35^ Our results suggest this leads to substantial retinal thinning in MS patients.

**Figure 5.**
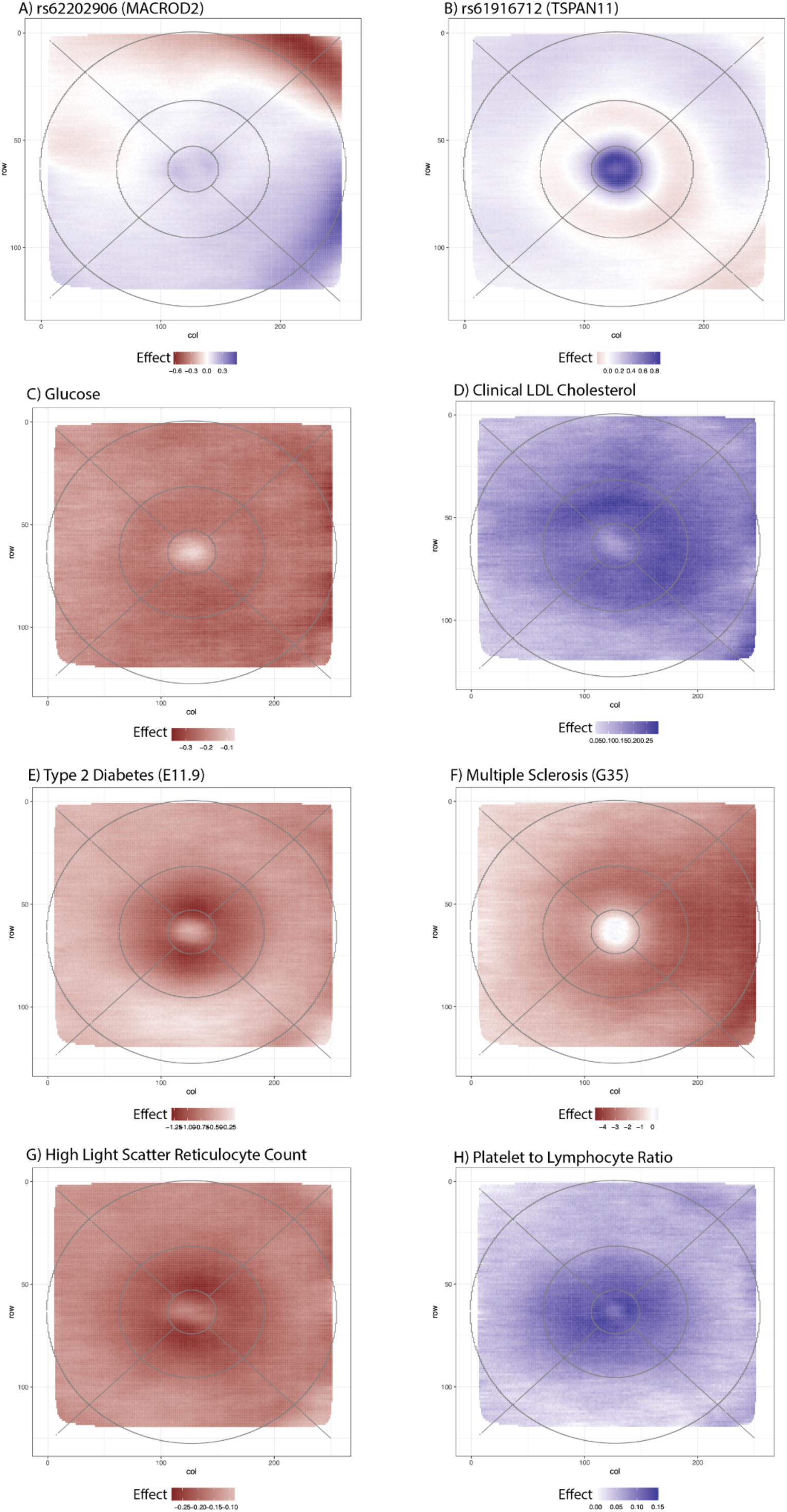
RT spatial effects of selected ‘omics signals. A: rs62202906 (*MACROD2*) B: Rs61916712 (*TSPAN11*), C: Glucose, D: Clinical LDL Cholesterol, E =Type 2 Diabetes w/o complications, F = Multiple Sclerosis, G: High light scatter reticulocyte count, H: platelet to lymphocyte ratio. Blue indicates a positive effect of the marker on retinal thickness while red indicates a thinning effect. In panels A-B effect size is change for each copy of the effect allele. For Panels E-F effect size is for presence of disease vs absence. In all other panels effect size is represented as 1 SD unit increase.

Metabolic and cardiovascular disorders were among diseases most strongly associated with thickness. Primary hypertension (“Essential (primary) hypertension”, ICD10-I10) displayed the highest amount of global significance, likely due to the high prevalence of this disorder (N=10,766). Type 2 Diabetes (T2D) without complication (“Without complication”, E11.9, N= 2,128, Figure 5F), pure hypercholesterolemia (E780, N= 5,227), and fatty liver (K76.0, N= 395) were also among the top RT-associated diagnoses. T2D and hypertension were also identified through our genetic correlation analyses, and are recognised comorbidities for multiple retinal disorders including AMD ^36^ and MacTel ^37^. T2D has a recognised retinal complication in diabetic retinopathy, affecting about one third of T2D patients, representing a major public health burden ^38^. These results complement our metabolic RT association results, where many of the metabolites most highly associated with RT have also been found to be associated with these disorders.

Investigation of retinal disorders was limited, given the study design of depletion of individuals with overt retinal disease. Nevertheless, we detected associations between RT and Cataract (H26, “Other cataract”, median pixel -log10 p-value = 4.49), and H25 “senile cataracts”,-log10 p-value = 2.38) as well as the “presence of an intraocular lens” (Z96.1, -log10 p-value = 3.51).

We also examined associations with broad comorbidity classes of ICD10 codes (Supplementary Table 18, Figure 4A), with “diabetes without chronic complication”, “renal disease”, “chronic pulmonary disease”, “mild liver disease”, and “peripheral vascular disease” showing the strongest associations with RT. Hierarchical clustering analysis revealed ten clusters of ICD10 diagnoses that comprised disease codes homogeneously affecting the thickness across the retina (Extended Data 9).

ORA revealed a pronounced susceptibility of the parafoveal ring to different diagnoses and diseases, confirming once again the sensitivity of this macular zone to disease (Figure 4C). Specifically, within this area of the retina, the effect is higher on the superior and temporal regions. Health insults on the parafoveal thickness were also highlighted by the Charlson comorbidity score (Supplementary Table 18), which presented higher scores associated with thinning of RT in this area.

We also examined associations with PRS for a number of diseases, as an alternative approach to ICD codes. This also allowed for investigation of associations between the disease risk rather than confirmed presence and also allowed investigation of a number of retinal diseases, and related phenotypes.

We examined 58 trait PRSs from 43,147 participants, and after correction for multiple testing, 48 PRSs were found to significantly affect the RT of at least one pixel (Supplementary Table 19). The averaged macular thickness ^8^ PRS was the most significantly associated, followed by retinal vascular caliber, AMD ^39^, and MacTel ^18^ (Supplementary Table 19). The more recent Han et al ^39^ AMD PRS resulted in a more powerful association signal than that released by the UKBB PRS release, based on Fritsche et al. ^40^

A higher burden of genomic risk of AMD, MacTel, T2D and celiac disease were all associated with parafoveal thinning. We found a significant negative association between RT and neuropsychiatric disorders such as Schizophrenia, Alzheimer’s and Parkinson’s disease although patterns of association differed across diseases. PRS related to metabolic traits (e.g., HDL cholesterol, celiac disease, HbA1c, apolipoprotein A1) as well as cardiovascular disorders (e.g. cardiovascular disease, resting heart rate, ischemic stroke, and atrial fibrillation) also showed significant association with thickness in particular areas of the retina (Supplementary Table 19).

We examined correlations of effects between the 25 diseases captured through both ICD codes and PRS in this study. We found the highest association agreement between ICD10 and PRS for type 1 and type 2 diabetes, asthma, Parkinson’s disease, as well as many cardiovascular disorders such as hypertension, thrombosis and strokes, coronary artery disease and atrial fibrillation (Supplementary Table 20).

### Associations with blood cell traits and markers of inflammation

After data pre-processing as detailed in Methods, data on 33 blood and inflammation biomarkers were able to be matched to RT data for 39,611 participants. Various reticulocyte traits were significantly associated with reduced retinal thickness, especially in the parafoveal area (Supplementary Table 21, Extended Data 10, Figure 5G), a finding consistent with our genetic correlation analyses.

Interestingly, higher counts of immune system cell types such as leukocytes and neutrophils were associated with parafoveal and temporal perifoveal thinning. Inflammation markers confirmed the high sensitivity of the parafovea to inflammation (Supplementary Table 21, Figure 5H), which has been previously reported for diseases such as Covid-19 ^41^.

We did not identify any association of RT with the 70 tested infectious disease antigens (Supplementary Table 22). This was likely due to the small sample size available for this analysis (n=764).

All association results at the pixel level are accessible through a user-friendly bespoke web browser (https://retinomics.org/).

## Discussion

Using one of the largest, systematically-collected OCT datasets in the world (UKBB), and through the application of AI, we generated the highest-resolution spatial dataset of RT ever produced, and demonstrated that the retina provides a window for human health. Our analysis reveals retina thickness to be exquisitely susceptible to a plethora of factors stemming from the genome, to metabolites, to blood traits, and diseases, with the parafoveal area most enriched for associations. Overall, we found reduced RT, or retinal thinning to be associated with poorer health, and increased burden of disease.

We employed two novel approaches to conduct genetic analyses using this data, firstly using dimensionality reduction, in the form of functional principal components, and secondly through our pixel-level approach, which required bespoke methods to summarise the data from over 29,000 GWAS. We note that a similar AI approach, applied to the UKBB whole body imaging data identifying genetic loci influencing skeletal proportions, some of which are risk factors for diseases of the skeleton, such as osteoarthritis ^42^. These complementary approaches, applied to AI-reprocessed data identified a greater number of loci, than previous GWAS conducted with RT derived directly from the TOPCON scanner, despite the heavy multiple testing burden. Moreover, these results give micron-level spatial resolution, whereas previous analyses were averaged across regions on the ETDRS, or Macula 6 grids. Many of our novel loci have previously been implicated in Mendelian retinal diseases, or ocular traits, and our GWAS findings overall were enriched for genes involved in eye development, cell differentiation and kidney development.

In some of our most important findings, reduced RT is highly associated with MS. This result provides strong, independent confirmation of multiple reports of the utility of OCT as the source of biomarkers for MS and MS progression ^43^, summarized in Britze et al ^44^. Additionally, it indicates that future studies utilizing RT as a biomarker should focus on the nasal perifoveal region of the macula as this region contains the greatest signal.

We additionally showed that a range of neurodegenerative and cardio-metabolic disorders were also highly associated with retinal thinning. We illustrate that the retina has unique metabolic sensitivities, with RT often affected in systemic metabolic diseases; indeed, metabolic disturbances that have already been identified in several retinal diseases ^18, 30, 45, 46^. For several of these metabolites and diseases, we supported these findings, through associations with directly measured phenotypes or their genetically-predicted proxies. We also identify that disease PRS sometimes reveal association signals not shown by their clinical record counterparts (ICD codes).

Further understanding of our genetic results will require single cell and spatial transcriptomics of the retina to investigate specific cell types being affected, as well as determining biological mechanisms.. Some of the genetic loci we describe here, and previously linked to disease, are already being investigated with these approaches.^17, 47^ Our work provides compelling evidence and extensive additional localising information to allow better targeting of the retina with these expensive technologies, as an increase of sample size will be required to investigate the common genetic variation that underpins much of the variation. Future work will extend this approach to retinal sub-layers which will further tease apart many of the associations since sub-layers are composed of particular cell types.

Our investigations focused on RT in retinal-normal individuals. In excluding individuals with overt retinal diseases, who may have major and unusual variation in RT, our analyses were undertaken on a more homogeneous cohort, thereby maximising our power for discovery. While our analyses primarily intended to capture variation in “healthy” eyes, it is likely some participants with mild retinal disease will have been included. Investigation of RT in the context of overt disease will also be of interest but will require a nuanced analysis approach which is beyond the scope of the current work.

This work has some shortcomings. The UKBB is very European-centric with >90% of participants being of European ancestries. Our cross-ancestry genetic association analyses highlighted significant genetic heterogeneity in RT, so investigation in individuals with greater ancestral diversity is vital; emerging efforts to increase diversity in population cohorts will hopefully allow this. The UKBB is a uniquely massive resource and as such we were only able to perform limited traditional validation of our multi-omics findings. However, we strove to validate many of the main findings through complementary approaches, such as utilising PRS alongside directly measured phenotypes. We note that whilst the UKBB is was intended to be representative of the aging UK population, it is subject to a range of selection biases; all described associations must be interpreted in this context.^48^

In summary, our work sets a new standard in multi-dimensional GWAS and ’Omics studies, supported by innovative statistical methods for imaging and GWAS at scale, but can only be fully appreciated through the bespoke web-browser that accompanies this work. This work validated and refined previous associations and will encourage others to reprocess high-dimensional data with AI. This work directs future biomarker studies and biological experiments.

## Methods

### Data and Data Access sharing

Data was sourced from the UK Biobank using project applications 28541. Research was approved by the Walter and Eliza Hall Institute of Medical Research Human Ethics Committee (HREC 17/09LR).

### OCT scans in UK Biobank

A subset of 85,726 UK Biobank individuals had retinal OCT data available. Scans were performed with the TOPCON 3D OCT 1000 Mk2. An OCT scan consists of a volume of 128 B-scans, each composed of 512 A-scans.

Scans were obtained on up to two occasions: “instance 0” (baseline); or ”instance 1” (first follow-up visit). The UK Biobank protocol states that one scan per eye should be recorded at each instance; however a subset of individuals had two scans for one or more eyes at a single instance, while some had only one eye measured at an instance. Initial assessment visits were conducted during 2006-2010, with a total of 68,530 participants. First repeat assessment visits were performed during 2012-2013, with a total of 19,579 participants. 2316 individuals had measures at both instances (Data-Fields 6070 and 6072 in UK Biobank).

At these visits, other relevant information collected included refractive error, intraocular pressure, visual acuity and eye checks with questions regarding any surgery that had been performed. In addition imaging device number was also recorded, which permitted technical error correction.

### Generation of macular retinal thickness data using machine learning

First, we developed a ML pipeline to extract the area between the internal limiting membrane (ILM) and retinal pigment epithelium (RPE) from OCT B-scans and to detect the layer boundaries without need for human annotations. The Topcon 1000 produces a 128 raster OCT volume over a 6mm*6mm*2.275mm area around the macula. The pixel resolution of each OCT B-scan is 512*650, and we measured the ILM-RPE thickness at every other pixel, i.e. 256 locations. The ML pipeline consisted of two steps. In the first step, we used the A star (A*) algorithm ^1^ to obtain layer boundaries by following the bright bands of the ILM and the RPE on OCT B-scans. These bright bands were then turned into segmentation masks for the ILM and RPE. A total of 6409 OCT-segmentation pairs were collected in this manner. These pairs were split 80% for training and 20% for validation at the subject level, and used as training data for a deep learning model. The deep learning segmentation model used was the Pyramid Parsing Network^2^ with a ResNet-18^3^ backbone. The model achieved a mean intersection over union (IOU) score = 0.97 on the validation set. After the model was trained, the layer boundaries were extracted by taking the top most pixel boundary for the ILM and bottom most pixel for the RPE in each predicted segmentation mask.

Next, we performed multiple rounds of quality control on the ML pipeline’s predicted segmentation masks. Quality control was performed at the location level with the location thickness rejected, and removed, if it failed any of the following six criteria. First, the thickness measurements from B-scans that were too faint to contain any ILM-RPE layers were removed (Supplemental Figure 1a). Second, thickness measurements for OCT scan regions that were too faint were removed (Supplemental Figure 1b). Third, locations where the location thickness was too thin (less than 30 pixels or 105μm) or too thick (more than 165 pixels or 577.5μm) were removed (Supplemental Figure 2a and 2b); for reference the typical thickness range is 154-232μm for males and 173-252μm for females. Fourth, regions where the predicted ILM or RPE boundaries were directly adjacent to regions that are too faint were removed as this suggests the OCT scan was not centred properly and was cut off (Supplemental Figure 3). Fifth, locations where the location thickness was discontinuous, i.e. where either the predicted ILM or the predicted RPE coordinates were discontinuous (Supplemental Figure 4). Sixth, the location thickness was rejected if the standard deviation of a rolling window of the 10 locations enclosing the location was very high (20 pixels or 70μm), i.e. highly unusual disturbances in the ILM-RPE layer (Supplemental Figure 5).

After the quality control, each OCT volume, containing 128*256 locations, was aligned to the fovea point. The fovea point was determined to be the centre of the area with the thinnest retina for each OCT volume scan and its location given as a tuple of slice number (between 1-128) and B-scan x-coordinate (between 1-256) giving a total of 32,768 measure locations for RT per eye. Moreover, the ILM-RPE thickness measurements for OCT scans from the left eye was reflected across the fovea point, so that it was aligned anatomically to the thickness measurements from OCT scans from right eyes.

**Hereafter, locations are referred to as pixels.**

### Quality control filtering of data

Additional filtering was conducted at the individual, OCT scan and pixel level for quality control. The overall filtering strategy is summarised in Supplementary Figure 8.

### Quality control filtering at individual participant or OCT scan level

The focus of the analysis was on healthy retina so individuals with overt retinal disease, or likely retinal disease were discarded (see Supplementary Table 1). Individuals with self-reported or mild, or non-retinal vision problems were retained, striking a balance between loss of power due to the removal of many participants and exclusion of some potential eye disease participants still being retained. Individuals with mild or non-retinal vision problems that were retained included those with: cataracts; reported as wearing glasses; myopia; astigmatism; presbyopia; hypermetropia. Additionally, participants were not excluded based on field 2227: “Do you have any other problems with your eyes or eyesight?”.

OCT scans were also removed based on Machine Learning model learnings. Quality control experiments as part of the machine learning modelling of the RT (described above) revealed diseased retinas (see examples in Supplemental Figure 6) for several individuals with no self-reported retina pathologies.

### Filtering of pixels, based on missingness

Scans were then trimmed to remove pixels with high levels of missingness or potentially spurious measurements. Firstly, pixels that were missing in >10% scans were removed. Then, entire rows or columns were removed if >50% of pixels had been removed based on the 10% missing threshold. Finally, pixel-level variance (across all scans) was estimated, and any pixel with variance > 55 was removed. This resulted in the removal of 3,727 pixels (11 %) and resulted in a trimmed grid of 29,041 pixels.

### Selection of scans and generation of pixel-level retinal thickness phenotypes

Using the trimmed grid, the best scan per eye, per instance, was selected for each individual. Firstly, scans were excluded if >10% of pixels in the trimmed scan were missing, or if refractive error for the corresponding eye and instance were unavailable. If there remained more than one scan available for an eye at an instance, the scan with the lowest missingness was retained, or in the case of a tie, one scan was randomly selected.

If an individual had scan data for both left and right eyes from the same instance, both eyes were selected. For the subset of individuals who had both eyes available at both instances, the instance with fewest missing data points (across both eyes) was identified, and the scans taken from that instance. For individuals who did not have a scan for both eyes measured at the same instance, the single eye with fewest missing data points was used (either eye/instance). In the case of ties, one eye was randomly selected.

### Imputation of missing values per scan

Imputation of scans was then carried out. For each scan, generalized additive model (GAM) was fitted using the “bam” function of mgcv R package (version 1.8-40) with the following model:

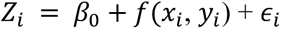

where *Z*_*i*_ is the depth measurement at position (*x*_*i*_, *y*_*i*_) and *y*(*x*_*i*_, *y*_*i*_) is a tensor product smooth function (k=12, 12). Where a pixel had missing data in an individual scan, the value was imputed using the predicted value from the GAM, assuming that missingness was at random.

### Final participant data generation

Finally the pixel-level retinal thickness phenotypes were generated using the imputed scans. If an individual had both left and right eyes available, the mean value for each pixel was taken, otherwise, the single available eye was used. A final filtering step removed individuals where their final scan phenotype had any pixels where thickness measurements were <30 or >150, or where the scan-wise standard deviation was > 15.

### Generation of fPC phenotypes

Using the final pixel-level retinal thickness phenotypes, functional principal component analysis (FPCA) was undertaken using the MFPCA R package (version 1.3-10)^4^, and using penalised 2D tensor product splines, with k=(12,12) as the maximum degrees of freedom. 100 FPCs were estimated, with the top six fPCs explaining >95% of the variance selected for follow-up analysis. For each individual, fPC scores were extracted, representing the contribution of each fPC to that individual’s scan phenotype.

### Quality control based on genetic data

The genotyping procedure, quality control and imputation of the UK Biobank cohort is described elsewhere.^5^ Imputed genotypes to the combined Haplotype Reference Consortium and UK10K panel (version 3; Category 100319) and genotype quality control metric files (Category 100313) were downloaded for the full UK Biobank cohort. Individuals were excluded if: they had been being excluded by UK Biobank, before imputation due to high heterozygosity or missingness (>5%); there was a mismatch between their genetically predicted and recorded sex, or they had a sex chromosome aneuploidy; they had an apparent excess number of relatives in the UK Biobank cohort (≥ 10 relatives); or they had withdrawn consent. Broad continental ancestries for all individuals were obtained from UK Biobank returned dataset 2442. Ancestry assignments were based on genetic similarity to individuals from the 1000 Genomes Project and Human Genome Diversity Panel, and were generated through the Pan-UK Biobank project (https://pan.ukbb.broadinstitute.org/). Individuals of European (EUR), Central and South Asian (CSA) and African (AFR) ancestries were identified for genetic analyses. The remaining individuals were excluded as their continental ancestry groups had <1000 individuals in each.

The full set of EUR, CSA and AFR individuals were then restricted to a maximally unrelated set. Pairwise relationships, up to 3rd degree were identified. Any individual with multiple (>1) relatives in the set were identified and removed. Remaining relative pairs were identified (each of whom were related to one individual in the set only). One of each pair was then randomly selected for exclusion.

Variants with minor allele frequency (MAF) < 0.05%, or imputation INFO scores < 0.8 were excluded. QCTool v2 was used for sample and variant based filtering, then bgen files were converted to PLINK binary format, using PLINK 2 (version 20221024).

### Genome-wide Association Analyses

Genome-wide associations were undertaken for the pixel-level retinal thickness phenotypes, and the fPC score phenotypes, using the glm command in PLINK 2 (version 20221024). Covariates included in the model were sex, age, age-squared, standing height, device ID, eye (left, right, or mean), spherical equivalent (for the corresponding eye, or the mean of left and right, if the mean of both eyes was used for the phenotype) and the first ten ancestry principal components as provided by the UK Biobank (field number 22009). Quantitative covariates were standardised to have mean 0 and covariance 1, and the variance inflation factor upper bound flag was set to 500, to allow for the multicollinearity of age and age squared. Chromosome X was analysed by coding hemizygous males as homozygous.

### Identification of independent significant loci

Identification of independent significant loci was carried out separately for the FPC analyses (6 GWAS) and the pixel-level analyses (29,041 GWAS).

Linkage disequilibrium (LD) clumping was undertaken for each set of GWAS results, using PLINK v1.90 to identify statistically independent signals, using strict criteria: index SNPs with p-value < 5E-8 identified, with clump SNPs having p-value < 5E-5, and r^2^ >0.001 and <5000kb distance to the index SNP. SNPs were allowed to belong to more than one clump. LD clumps were collated to give a list comprising all genome-wide significant (p < 5E-8) index SNPs, and the corresponding clump SNPs. The results for all N SNPs in this list were then extracted from each GWAS (N_pixel-levelSNPs_ x 29,041 associations for the pixel-level analyses; N_FPCSNPs_ x 6 associations for the FPC analyses).

Identification of loci was then carried out, using the extracted associations, and the clumping results, here, described in the context of the pixel-level results:

While at least one pixel-SNP association has p-value < 5E-8:

1. Order all extracted associations by P-value.
2. Take the pixel-SNP combination with smallest association p-value.
3. If this SNP is an index SNP in the clumping results for this pixel, then:

a. Define a new locus with the index SNP as the sentinel SNP and the pixel as the top pixel for the locus.
b. Find all corresponding clump SNPs (P < 5E-5) for that locus, from the clumping results; these SNPs are allocated to the locus as supporting SNPs.
c. Find all pixels with P < 5E-5 for either the sentinel, or any of the supporting SNPs; these pixels are allocated to the locus.
d. Remove all allocated SNP-pixel combinations from the list of extracted associations.
4. Else, if this SNP is not an index SNP in the clumping results for this pixel, then:

a. Find the clump where the SNP is included as a clump SNP.
b. Extract the index SNP for this clump, and confirm this index SNP has already been assigned to a locus.
c. Remove any remaining clump SNPs from the list of extracted associations.
5. Repeat steps 1-4 until no pixel-SNP associations with p-value < 5E-8 remain.

Loci were identified from the 6 FPC GWAS, using an equivalent approach.

The above process resulted in two lists of loci: one for the pixel-level analyses, containing loci with at least one pixel-SNP association meeting P < 5E-8; one for the FPC analyses, containing loci with at least one FPC-SNP association meeting P < 5E-8. Finally, these lists were filtered, to remove loci with fewer than 5 SNPs assigned (ie fewer than 5 SNPs in the region with p < 5E-5), as these were deemed to be likely false positives.

### Reporting of GWAS loci

The loci identification process utilised the conventional genome-wide significance level (P < 5E-8), however this does not account for the heavy multiple testing burden of our approaches. The final reporting, and subsequent follow-up of loci was therefore restricted to SNP associations, meeting conservative Bonferroni corrected thresholds: P<5E-8 / 29,041 = 1.72E-12 for the pixel-level analyses; P <5E-8 / 6 = 8.33E-9 for the FPC analyses.

For completeness, we provide the unrestricted loci lists (P < 5E-8) in the supplementary results, with loci meeting the strict Bonferroni thresholds flagged (Supplementary Tables 5 and 6).

Sentinel SNPs were annotated using the variant effect predictor (VEP v.109, ^6^). Loci were also annotated to indicate whether they were identified through previous GWAS of retinal thickness ^7–9^. For these annotations, the sentinel SNP along with all other SNPs allocated to each locus were compared to previously reported SNPs. If no SNP allocated to a locus was included amongst previously reported SNPs, that locus was deemed novel.

### Follow-up of GWAS loci

Follow-up was undertaken for loci meeting the strict Bonferroni corrected thresholds described above. Mapping of loci to candidate genes via positional mapping, eQTL data, and chromatin interaction data, was undertaken based on annotations obtained via FUMA (v1.5.3).^10^ For each set of loci (pixel-level associated loci; fPC associated loci), the list of sentinel SNPs was uploaded as a predefined list of lead-SNPs, with mapping of SNPs to genes carried out using the uploaded sentinels, and all SNPs in LD with r2 > 0.5 with the sentinels (collectively referred to herein as candidate SNPs).

Positional mapping identified genes overlapping with, or <=10kb from, candidate SNPs using ANNOVAR (2017-07-17). eQTL mapping identified genes (cis, up to 1Mb) whose expression are associated (FDR<0.05) with candidate SNPs, in retina (EyeGEx)^11^, blood (GTEx v8 tissues: EBV-transformed lymphocytes; whole blood) or brain (GTEx v8 tissues: amygdala; anterior cingulate cortex BA24; caudate basal ganglia; cerebellar hemisphere; cerebellum; cortex; frontal cortex; hippocampus; brain hypothalamus; nucleus accumbens basal ganglia; putamen basal ganglia; Spinal cord cervical c-1; brain substantia nigra).^12^ Chromatin interaction mapping used HiC data from fetal and adult human brain, to link candidate SNPs to genes based on overlapping enhancer-promoter and promoter-promoter interaction regions (restricted to significant interactions with P < 2.31e-11).^13^

The candidate genes identified through these mappings, were additionally annotated as to whether they were associated with eye or retina-related phenotypes in mice or humans. For mouse phenotypes, we utilised a set of genes from the International Mouse Phenotyping consortium (https://www.mousephenotype.org), causing “abnormal eye morphology” (<u>MP:0002092</u>), and with human orhologs.^14^ To identify relevant genes in humans, we queried the OMIM database for genes with “retina” or “retinal” included in the entry, and clinical synopsis including “Head & neck”.^15^

For completeness, we report all genes implicated via the above mappings, or annotations (Supplementary table 10). For certain follow-up analyses, we further prioritised candidate genes, based on the number of lines of evidence for that gene (prioritisations specified with the relevant analyses). To this end, we created a score, which summed up whether the gene was: i) implicated via positional mapping; ii) implicated via mapping with an exonic candidate SNP; iii) implicated via mapping with a candidate SNP with a CADD score >= 20; iv) implicated via retina eQTL data; v) implicated via blood eQTL data; vi) implicated via brain eQTL data; vii) implicated via HiC data from brain; viii) associated with eye abnormalities in mice; ix) associated with a retina OMIM phenotype. This score is reported in the “evidenceScore” column of Supplementary Table 10.

### Clustering of sentinel SNP effects

We performed unsupervised hierarchical clustering analysis on both pixels and SNPs, based on the effect sizes for the sentinel SNPs, across all 29,041 pixels. The number of clusters was determined from visual inspection of the clustering dendogram.

### Associations in non-European ancestries

We undertook association analyses for all identified loci, in individuals from UK Biobank AFR and CSA ancestries. For each locus, we examined associations with the sentinel SNP at each identified locus with all 29,041 pixels (pixel-level analyses), or for the FPC(s) for which the SNP was significant. Association analyses was undertaken using PLINK 2 (version 20221024), similarly to the analyses of European individuals. For each locus we report the results for the pixel, or FPC, with the most significant association observed in European individuals. We examine concordance of direction of effect across ancestries. We deemed associations in AFR and CSA individuals to be statistically significant, if they met the Bonferroni corrected p-value threshold for the number of loci tested. We additionally compared whether the pattern of association across the entire scan was similar across ancestries. For each SNP, we calculated the scan-wise correlation (i.e., the correlation of effects across all pixels), using Spearman’s Rank Correlation.

### Genetic correlation analyses

Genetic correlation analyses were carried out for fPCs 1-6, using LD-score regression^16^, implemented via the Complex Traits Genetics Virtual Lab.^17^ We examined correlations with 1309 traits in total (Supplementary Table 12).

### Gene set over-representation analysis

Gene set over-representation analysis (ORA) for Gene Ontology Terms was performed using the R-package *ClusterProfiler (v. 4.2.2).*^18^ ORA was carried out for genes implicated via the pixel-level and fPC results separately, using two sets of genes in each case: (i) all candidate genes (ii) the gene(s) with the most lines of evidence (i.e. highest “evidenceScore”) for each locus. Correction for multiple testing was performed using Benjamini-Hochberg ad-hoc correction, and we report all GO terms with corrected p-value < 0.05.

### Other ‘omics analyses

We next assessed the associations with metabolomics, blood and immune trait markers, and disease codes (ICD codes) on RT. For metabolite and disease codes, we examined associations with both direct measures, and using polygenic risk scores. These analyses were restricted to EUR individuals, due to limited sample sizes for other ancestries. We outline the cleaning of each data type, then the analyses undertaken,

### Polygenic risk scores preparation

Polygenic Risk Scores (PRS) were used to investigate relationships of traits with RT. We used three types of PRS in this study. Type 1 PRS comprise *trait PRS* composed mainly of PRS describing disease susceptibility. Type 2 PRS comprise *metabolic PRS* measuring genetic predisposition to blood abundance of a set of metabolites. The *trait PRS* are further subdivided into three subclasses. Two of these, the *Standard* and *Enhanced PRSs*, were downloaded from the UK Biobank having been generated and deposited in the UKBiobank as described by Thompson and colleagues ^19^ (Categories 301 and 302). These PRS had already undergone extensive QC and data processing, thus were used, as is, in the association analyses. The third subclass of the trait PRS are defined as *Internal PRS (*Type 3 PRS). These were PRS of particular relevance to the RT phenotype and included two macular disorders and two measures related to the RT phenotype. This included PRS for: (i) Macular Telangiectasia type 2 ^20^, (ii) Age-related macular degeneration ^21^, (iii) Retinal thickness ^7^, and (iv) retinal venular and arteriolar calibre ^22, 23^. To construct these PRS we extracted the top SNPs at each genome-wide significant locus associated with the trait as reported by the authors. We then used the R-package *bigSNPR* (*version 1.12.2*)^24^ to extract the selected SNPs from UK Biobank and create the polygenic risk scores.

*Metabolic PRSs* were defined using metabolites measured with the Biocrates platform which was used previously to determine genetic associations ^25^. Similar to the *Internal trait PRSs* we selected the SNPs for each *metabolite PRS* as the top associated SNP for each locus. We then discarded all metabolites that had fewer than three top SNPs available to construct the PRS, as measured in the UK Biobank. We then used *bigSNPR* to combine the SNPs into a single PRS for each metabolite.

### Blood traits data preparation

Blood trait data (Category 100081) was downloaded from UK Biobank. Blood trait data was collected using Beckman Coulter LH750 instruments for all 500,000 participants of the UK Biobank for the baseline visit. Cleaning of the blood traits was undertaken, based on the methods employed in ^26^. In brief, all blood traits were first log, or logit (in the case of ratios or proportions) transformed. Using a restricted set of central measurements (measures <3.5 median absolute deviations from the median), a generalised additive model (GAM, using the mgcv version 1.8-40 R package) was fitted, to model the effects of several technical covariates (time of measurement, instrument, acquisition route, day of the week). In the full dataset, model residuals were calculated and used to generate traits adjusted for technical effects. Finally, outliers (>6 median absolute deviations from the median) were excluded. The log/logit transformed, adjusted traits were used for downstream analyses.

Following the work from Nøst and colleagues ^27^ we used this data to calculate systemic inflammation markers. Specifically, we used peripheral lymphocyte, neutrophil, monocyte and platelet counts to calculate neutrophil-to-lymphocyte ratio (NLR), platelet- to-lymphocyte ratio (PLR), lymphocyte-to-monocyte ratio (LMR) and the systemic immune-inflammation index (SII). These were analysed in the same manner as the directly measured blood traits.

### Infectious disease antigens preparation

Infectious disease antigen data (Category 1307) was downloaded from UK Biobank. Antigens were measured using the Luminex platform. Only samples whose antigens were measured were kept in the dataset. Antigen measurements were log transformed prior to use.

### Metabolomics data preparation

Measurements of metabolic data from non-fasting venous blood of around ∼120,000 UK Biobank participants (Julkunen et al. 2021) were downloaded (Category 220). Metabolic quantifications included a total of 249 metabolic biomarkers. Of these, 168 were directly measured and quantified by NMR spectroscopy using the NIghtingale platform, while 81 biomarkers were ratios or derivative measurements. The metabolomics data was processed with the R package *ukbnmr (version 1.5)* ^28^. This procedure removed technical variation as well as normalising the metabolic values. Non-derived metabolites were then square-root transformed to achieve symmetry of the distribution. Assuming missingness at random, metabolic values were imputed using the Multiple Imputation Chain Equation by the R-package *MICE (version 3.15.0)* ^29^. Five different imputations of the entire data were generated and the average of these five imputation set values was retained as the final set of imputation values. Composite metabolites and ratios were then re-computed using *ukbnmr*. A total number of 325 metabolites were included in this study.

### Disease ICD10 codes and Charlson comorbidity scores data preparation

To extract ICD10 diagnoses for each individual we used the R-package *ukbtool (version 0.11.3)* (Hanscombe et al. 2019). This data was then transformed into binary format where each patient was represented either by 0 or 1 for each ICD10 code depending on whether they were unaffected or affected. Additionally, using the R-package *icd (version 4.0.9)* (https://github.com/jackwasey/icd) we constructed 17 comorbidity scores as defined by Quan and colleagues (Quan et al. 2005). Using the same tool we additionally constructed a Charlson Comorbidity Index (CCI) for each individual which is calculated using the inpatient’s disease category, obtained from the diagnosis made during hospital admission. Each of the 17 scores is weighted with a value of 1-6 and then all comorbidities are summed together.

### -Omics Data Analyses

PRSs, metabolomics, ICD10 scores, CCI scores, blood traits and infection antigens were tested against retinal thickness using the same protocol. Firstly, all data from participants from any of these omics where no imaging data was available was discarded. Secondly, to allow for comparison of effect sizes across biomarkers/phenomes, each of them was standardised to have mean equal to zero and standard deviation equal to one. Thirdly, association testing was performed by regressing each marker against all retinal pixels thicknesses correcting for the following covariates: (i) genetically derived sex at birth, (ii) age, (iii) imaging device number, (iv) standing height, (v) mean refractive error measured by spherical equivalent, (vi) eye and (vii) the first ten genetic principal components. Participant data presenting with missing information for any of these covariates was discarded. Regression analysis was performed using the package *limma (version 3.50.3)* ^30^ which applies an empirical Bayes framework to model relationships between outcome variables (here RT pixels). Fourthly, multiple testing correction on p-values was performed using the ad-hoc R-function *p.adjust*. All p-values within each omic type were corrected jointly using the Benjamini-Hochberg correction for false discovery rate.

The same strategy as described above was used to test each association of each omics with the first six fPCs. Given that the fPCs capture specific grid-wide patterns, a more lenient FDR threshold of 1% was used to define significance of this analysis.

Interaction effects between metabolic levels and age on retinal thickness were evaluated following the same approach as that described above, but by adding an interaction term for metabolic level and age, in addition to the main effects, in the limma linear regression model. To test whether the effect of the metabolites on thickness varied with age, we counted the proportion of significant interaction terms where the sign is equal to the sign of the main effect of the metabolite on retinal thickness.

Correlation between PRS and ICD10 pixel-level effects was performed by simply calculating the Pearson’s correlation coefficient across all pixels by matching PRS disease effect with the available ICD10 codes. For each pair, three analysis were performed, one using every pixel, one using only those pixels where the PRS had a significant effect, and one using only those pixels where both the PRS and ICD10 had significant effects.

For plotting and interpretation purposes, we summarised each biomarker’s effect on global retinal thickness using a suite of measures. These were: (i) number of pixels significantly affected by each biomarker, (ii) the average beta of the biomarker across all pixels, (iii) the median -log10(p-value) of the marker association with pixels, (iv) Values for all of these measures are available as columns in each of the associated result tables, which are specific for the eight types of variables being tested (Supplementary Tables 14-22). Each results table also has biomarker specific descriptors in the columns preceding these measures.

We performed hierarchical clustering analysis on both pixels and biomarkers within the snps, metabolomics and blood traits analyses. These omics were selected for this analysis due to their large number of biomarkers and prior knowledge of the existence of structured correlation patterns. Clustering of biomarkers was performed to capture shared (spatial) effect patterns on pixel-level RT. Clustering was performed using unsupervised hierarchical clustering. The number of clusters was determined visually based on the hierarchical clustering tree produced by the analysis.

Pixel-level over-representation analysis (ORA) was performed by using the R-package ClusterProfiler (*version 4.2.2*) ^18^. Each pixel *i* was defined as a set and each test pixel *i*-marker was defined as an entry of the set. Over-representation of significant pixel-marker association between pixels was then evaluated. ORA was only performed for the metabolomics biomarker main effect results, the metabolite-age interaction results and the ICD10 diagnosis given. For ICD10 codes ORA was performed on a random grid of pixels due to the computational burden. Plotting of ORA results was performed by smoothing log10(p-values) over the 2D grid using the loess function.

### Display of association results with RT (pixel level)

All association results are displayed using the same orientation for results as per Supplementary Figure 1.

Top = superior, towards top of head

Bottom = inferior, towards bottom of head

Left hand side = temporal, towards the ears

Right hand side = nasal, towards the nose

Results are displayed either as p-values (adjusted or unadjusted), which may be transformed with a log10 transform, or as betas (effect of the biomarker/phenotype), or a transform of beta.

The Early Treatment Diabetic Retinopathy (ETDRS) grid,^32^ which segments the macula into nine sectors and is widely used, is overlaid on most images to provide further spatial context for the interpretation. This grid was added by drawing the grid coordinates onto the each plot and thus represents an approximate mapping. (Supplementary Figure 1)

### Web Browser

An interactive website, https://retinomics.org, was developed to help visualize the GWAS and metabolomics results in the paper. The website has interactive pages for each type of biomarker of interest, such as GWAS, metabolites, hematology, infections, polygenic risk scores and ICD10 codes. For each type of biomarker, there are two views. First, there are heatmap plots to show the effect and pvalue of selected biomarker of interest. Second, there are macula location plots that shows the link between all significant biomarkers for that macula location. The location plots are manhattan/region plots for the GWAS and volcano and bar plots for the other types of biomarkers. Users can interact with the plots by clicking or selecting a biomarker of interest to trigger the loading of its heatmaps or click a macula location in the heatmaps to load its corresponding location plots. The website was implemented in HTML, native Javascript, and d3js.

All results are displayed with the same orientation on the pixel grid as described in Supplementary Figure 1.

## Data Availability

All data used in this study can be accessed from the UK Biobank upon request, via an application process (https://www.ukbiobank.ac.uk/).

RT pixel-level and the six 2D fPC data will be returned to the UK Biobank.

GWAS summary statistics for the six 2D fPCs will be submitted to the GWAS catalogue. Other ‘Omics summary statistics will be available for download at https://retinomics.org/.

## Code Availability

We used publicly available open-source software for these analyses. Scripts for the genetic analyses can be found at https://github.com/bahlolab/retinalThicknessGWAS

## Acknowledgements

This work was supported by funding from the Australian Government National Health and Medical Research Council (NHMRC) from APP1195236 (M.B.) and APP1181010(B.R.E.A.).

This work was also made possible through the Victorian State Government Operational Infrastructure Support and NHMRC Independent Research Institute Infrastructure Support Scheme (IRIISS). We also thank the Lowy Medical Research Institute (LMRI) for laboratory support to A.Y.L. and M.B. and institutional support to C.Y.E.. C.Y.E. and A.T.l received a proportion of their financial support from the UK Department of Health through an award made by the National Institute for Health Research to Moorfields Eye Hospital NHS Foundation Trust and UCL Institute of Ophthalmology for a Biomedical Research Centre for Ophthalmology. A.T. and C.Y.E. acknowledge financial support from the Said Foundation.

## Author Contribution Statement

M.B. and A.Y.L. conceptualised the study and co-led the project. C.E., A.T. and K.W. shared clinical expertise to remove retinal disease individuals and other confounders from the UKBB cohort used in the primary analysis. A.Y.L. led the development of the CNN for the OCT data. Y.W., supported by J.O. and Y.K., developed the CNN. Y.W. developed the webportal with feedback from R.B. and overall direction from A.Y. and M.B.. V.E.J. and R.B. conducted association analyses, generated figures and results. B.R.E.A. provided feedback on analyses and figures. S.F. developed early analyses for the genomics data. M.L.G. contributed interpretation of the metabolomics results. M.B. drafted the manuscript and led the ‘omics analyses. All authors read and provided feedback on the manuscript.

## Materials and Correspondence

Correspondence and requests for materials should be addressed to Melanie Bahlo.

## Supplementary information

SupplementaryInformation.pdf contains Extended Data 1-10 and Supplementary Figures 1-15. SupplementaryTables.xlsx contains Supplementary Tables 1-22.

