## Supplementary Information for "Multi-omic spatial effects on high-resolution AI-derived retinal thickness"

#### Extended Data Index

**Extended Data 1**. Retinal Thickness fPC representations.
**Extended Data 2:** Summary of RT measures.
**Extended Data 3.** Summary of genetic loci.
**Extended Data 4.** Manhattan plots of GWAS for fPCs 1-6 (A-F).
**Extended Data 5.** Cross ancestry comparisons
**Extended Data 6.** Genetic Correlations.
**Extended Data 7.** Metabolomics results.
**Extended Data 8.** Polygenic risk scores results.
**Extended Data 9.** Clustering based on ICD10 results
**Extended Data 10.** Blood and inflammation markers results.

#### Supplementary Figure Index

**Supplementary Figure 1.** Quality exclusion criterion 1 - faint B-scans.
**Supplementary Figure 2.** Quality exclusion criterion 2 - Thickness measurements too thin in scan region.
**Supplementary Figure 3.** Quality exclusion criterion 3 - Thickness too thin at specific locations**.**
**Supplementary Figure 4.** Quality exclusion criterion 4 - Discontinuities in ILM-RPE segmentation.
**Supplementary Figure 5.** Quality exclusion criterion 5 - Location thickness discontinuities.
**Supplementary Figure 6.** Quality exclusion criterion 6 - undocumented retinal disease.
**Supplementary Figure 7.** Early treatment of diabetic retinopathy (ETDRS) grid with orientation used for all retinal images.
**Supplementary Figure 8.** Quality control filtering.
**Supplementary Figure 9.** Two dimensional functional principal component analysis.
**Supplementary Figure 10:** Pixel-level associations with basic characteristics.
**Supplementary Figure 11.** Procedure for collation of pixel-level GWAS results.
**Supplementary Figure 12.** Comparison with previously identified GWAS loci.
**Supplementary Figure 13.** Clustering of SNPs identified through the pixel-level analyses, based on effects across all 29,041 pixels.
**Supplementary Figure 14.** Clustering of pixels, based on SNP effects for loci identified through the pixel-level analyses.
**Supplementary Figure 15.** Gene Ontology Over-Representation Analysis

### Supplementary Table Index

**Table Overview.** Overview of Tables
**Column Descriptors A**. Column descriptors for Supplementary Tables 5-12
**Column Descriptors B**. Column descriptors for Supplementary Tables 13-21
**Supplementary Table 1**. Available OCT data, and participant exclusions
**Supplementary Table 2**. Basic characteristics for the analysed cohort
**Supplementary Table 3**. Associations between RT fPCs and basic characteristics
**Supplementary Table 4.** Sample sizes for -omic association analyses
**Supplementary Table 5.** Loci identified through pixel-level GWAS
**Supplementary Table 6.** Loci identified through fPC GWAS
**Supplementary Table 7.** Positional mapping
**Supplementary Table 8.** eQTL mapping
**Supplementary Table 9.** Chromatin interaction mapping
**Supplementary Table 10.** Summary of GWAS signal to gene mapping
**Supplementary Table 11.** Cross-ancestry comparison of genetic effects
**Supplementary Table 12.** Gene Ontology Over-representation analyses
**Supplementary Table 13.** Genetic Correlations
**Supplementary Table 14.** Metabolite traits association analysis with retinal thickness
**Supplementary Table 15.** Metabolite traits interaction analysis with retinal thickness
**Supplementary Table 16.** Metabolic polygenic trait association analysis with retinal thicknes
**Supplementary Table 17.** ICD10 code association analyses with retinal thickness
**Supplementary Table 18.** Co-morbidity association analyses with retinal thickness
**Supplementary Table 19.** Polygenic traits association analysis with retinal thickness
**Supplementary Table 20.** Corelation between ICD10 codes and respective PRS
**Supplementary Table 21.** Blood and Inflammation marker association analyses with retinal thickness
**Supplementary Table 22.** Antigen association analyses with retinal thickness

### Extended Data

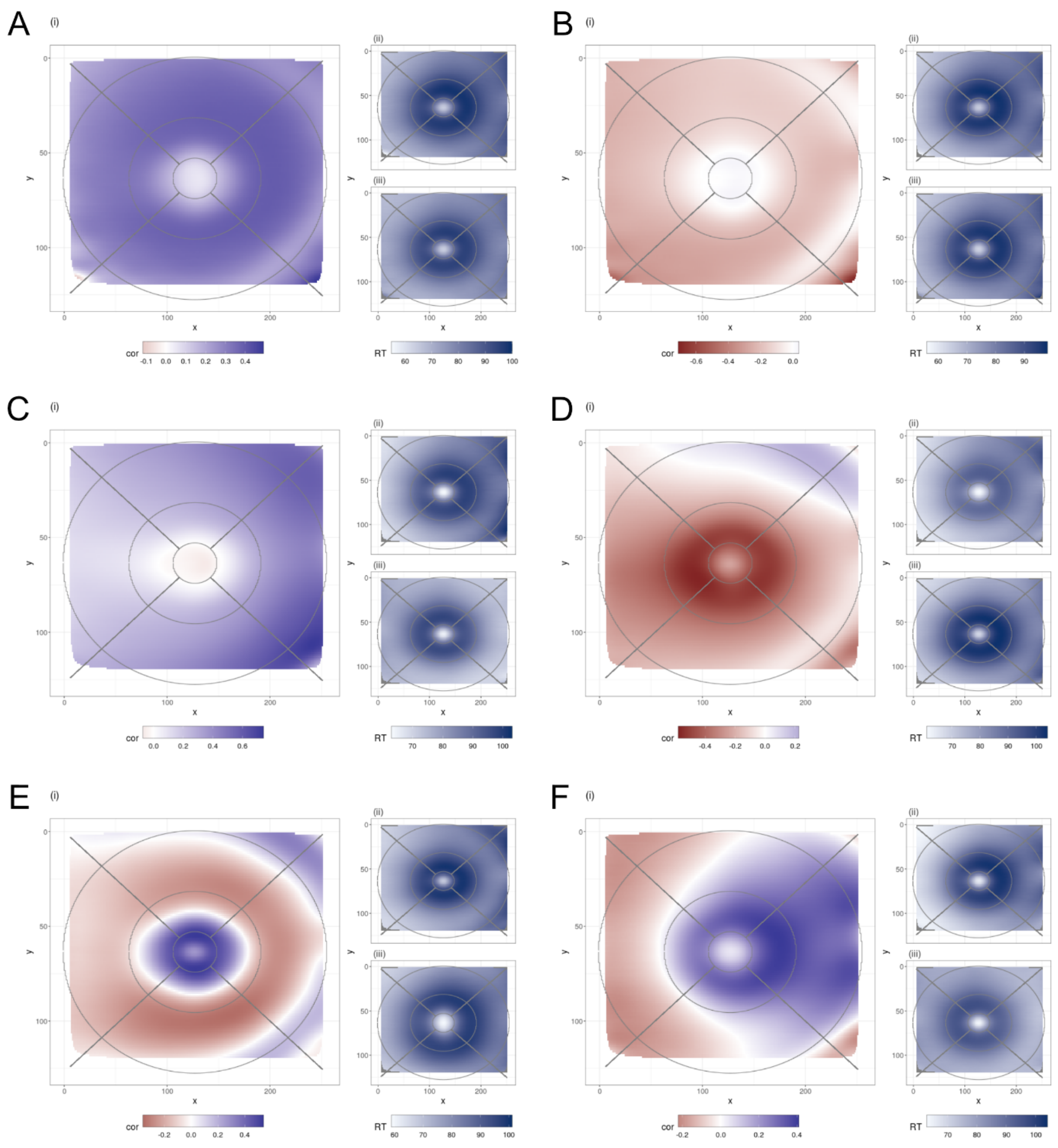

**Extended Data 1. Retinal Thickness fPC representations.**
For each fPC 1-6 (A-F): (i) shows the correlation between individual fPC scores and each pixel wise RT value. Higher absolute correlations indicate a greater contribution to the fPC for that pixel. (ii) and (iii) show the averaged RT scan for the 100 individuals with the (ii) highest and (iii) lowest score values for that fPC, giving an idea of the dynamic range for the RT spatial patterning captured by each fPC.

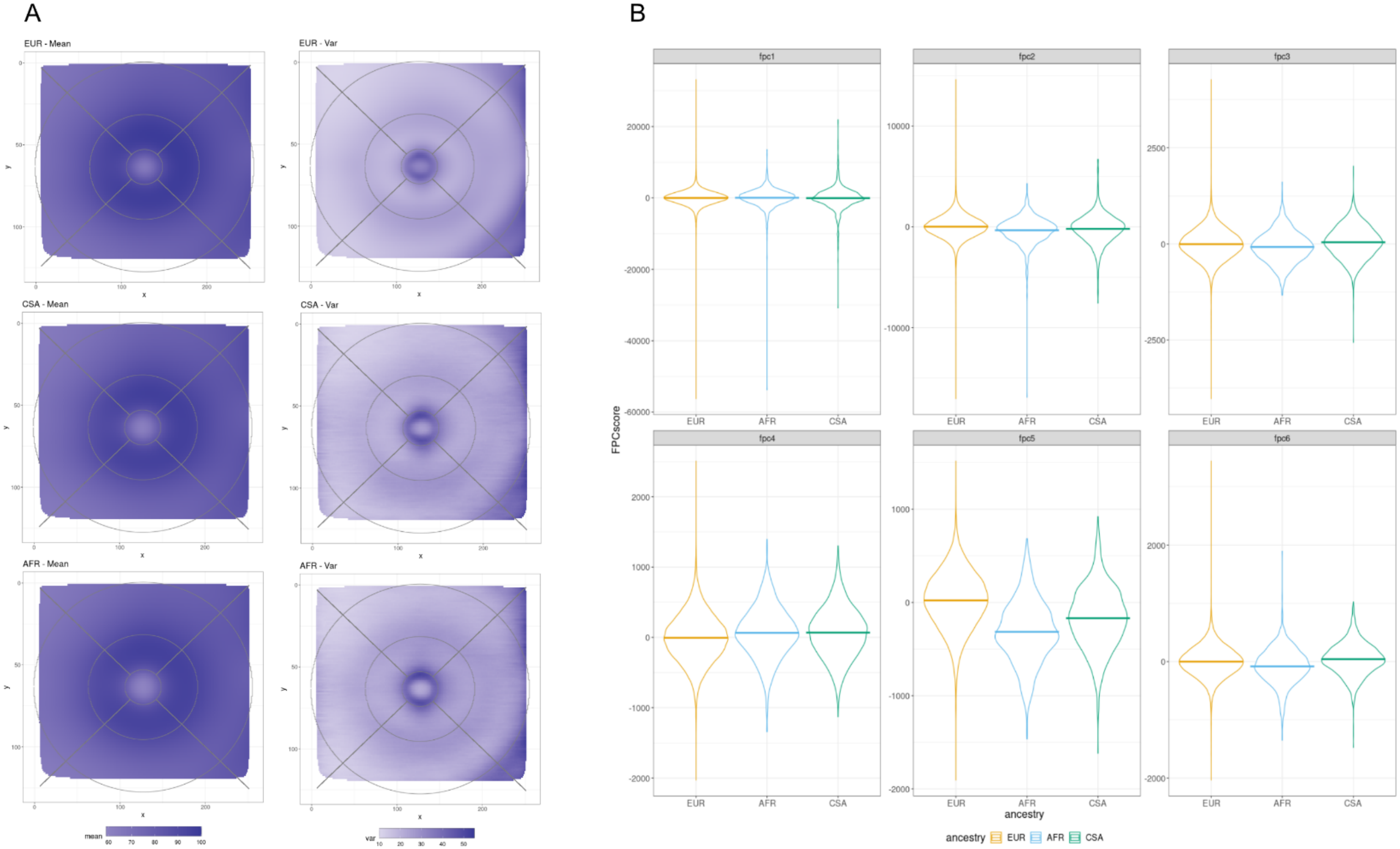


**Extended Data 2: Summary of RT measures.** Summary of A. pixel-level, and B. fPCs of RT measures, stratified by ancestry.

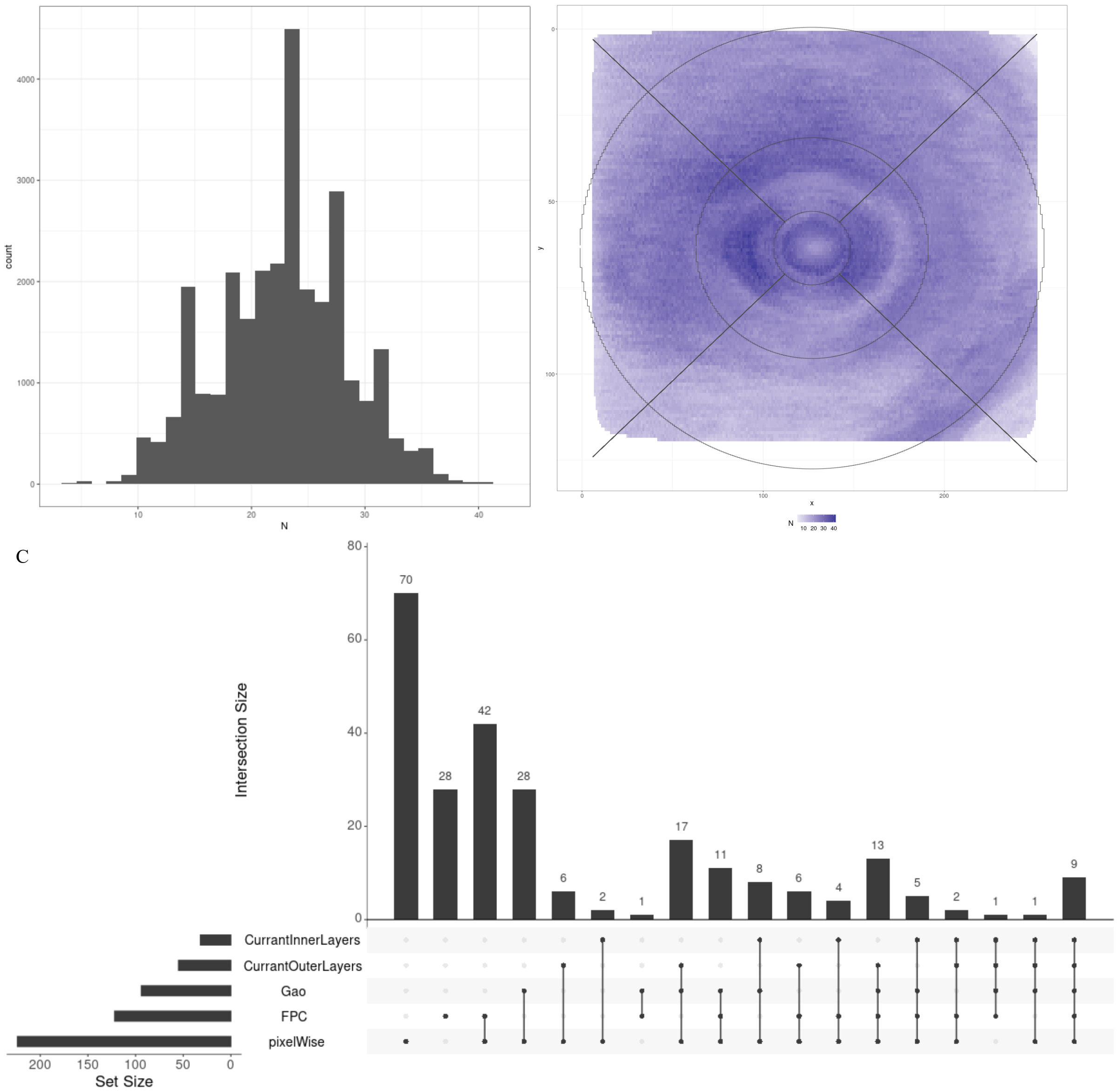


**Extended Data 3. Summary of genetic loci.** (A) Histogram and (B) spatial distribution, of the number of Bonferroni-corrected significant loci (p<5e-08/29,041) for the pixel-level GWAS results. (C) Comparison of loci identified through the pixel-level and/or fPC analyses, with loci identified in previous GWAS of RT measures (Currant et al. 2023; Currant et al. 2021) and(Gao et al. 2018), displayed as an upsetR plot. For these annotations, the sentinel SNP along with all other SNPs allocated to each locus were compared to previously reported SNPs. Plot only includes loci identified in the present analyses.

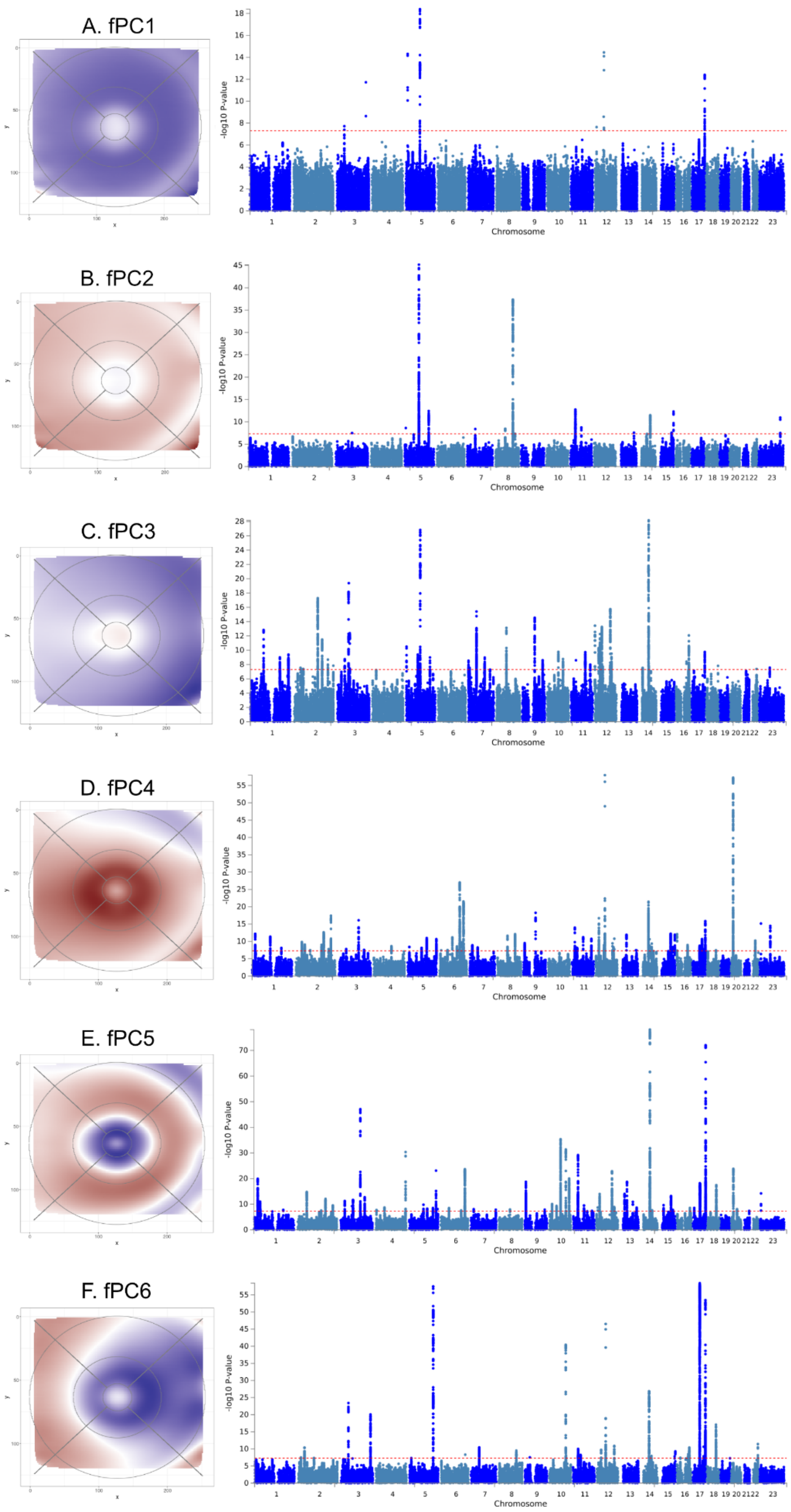


**Extended Data 4**. Manhattan plots of GWAS for fPCs 1-6 (A-F).

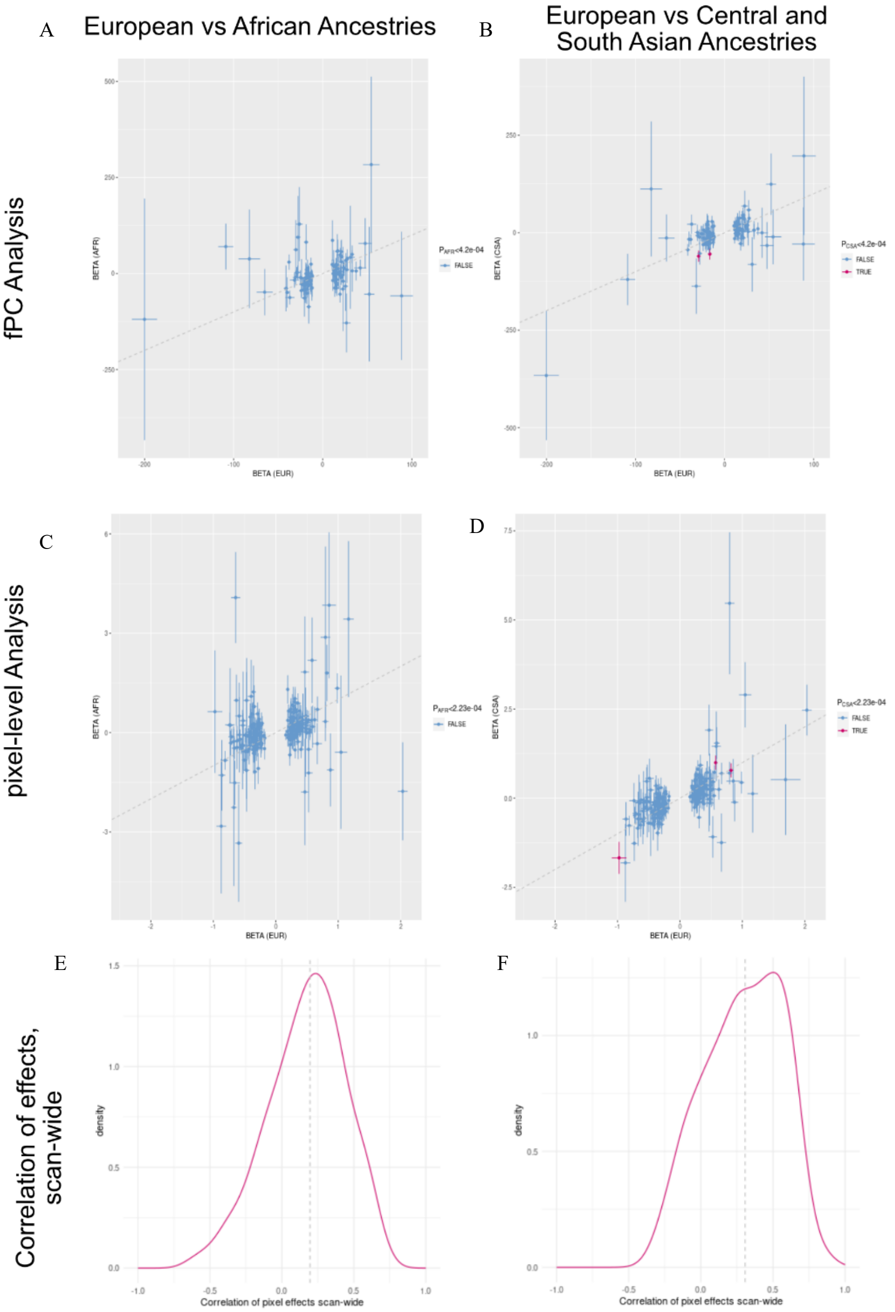


**Extended Data 5. Cross ancestry comparisons.** Cross ancestry genetic effect size comparisons of effect estimates for EUR vs AFR (A, C, E) and EUR vs CSA (B, D, F). A-D show comparisons of effect sizes (betas), with standard errors, from genetic association analyses for the sentinel SNP and the top pixel/fPC, for all loci identified through RT pixel-level (C and D) and fPC (A and B) analyses detected in the EUR discovery analyses. Loci meeting the Bonferroni corrected significance threshold in the AFR/CSA individuals for each analysis indicated in red. E and F show the distribution of the scan-wide correlations of effect sizes (ie the correlation of effects across all pixels), for each sentinel SNPs identified through the pixel-level analysis. The dotted line shows the median scan-wide correlation.

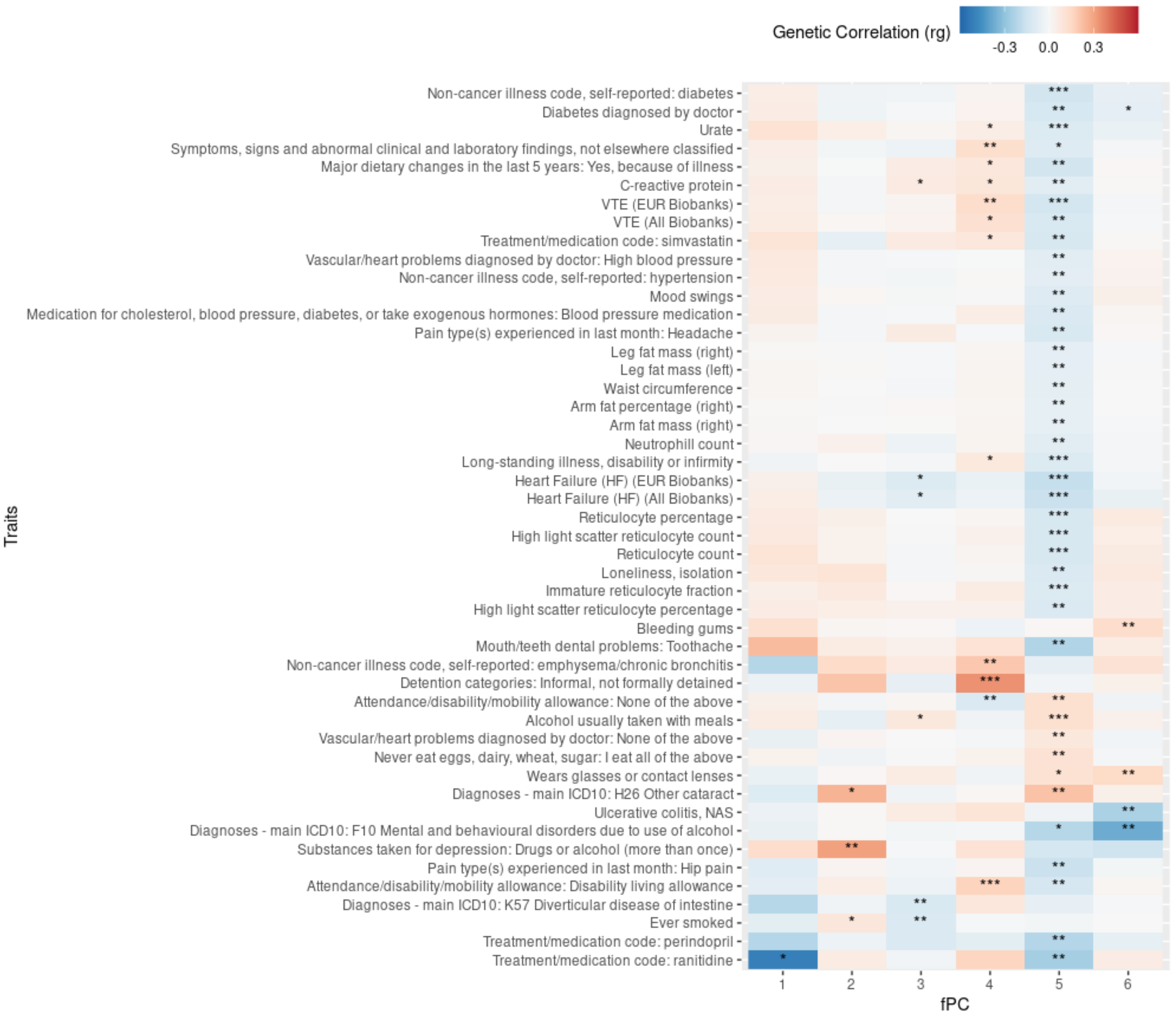


**Extended Data 6. Genetic Correlations**. Traits showing the most statistically significant genetic correlations with RT fPCs 1-6. Significance of genetic correlation: * P < 0.05; ** P < 0.005; *** P < 0.0005).

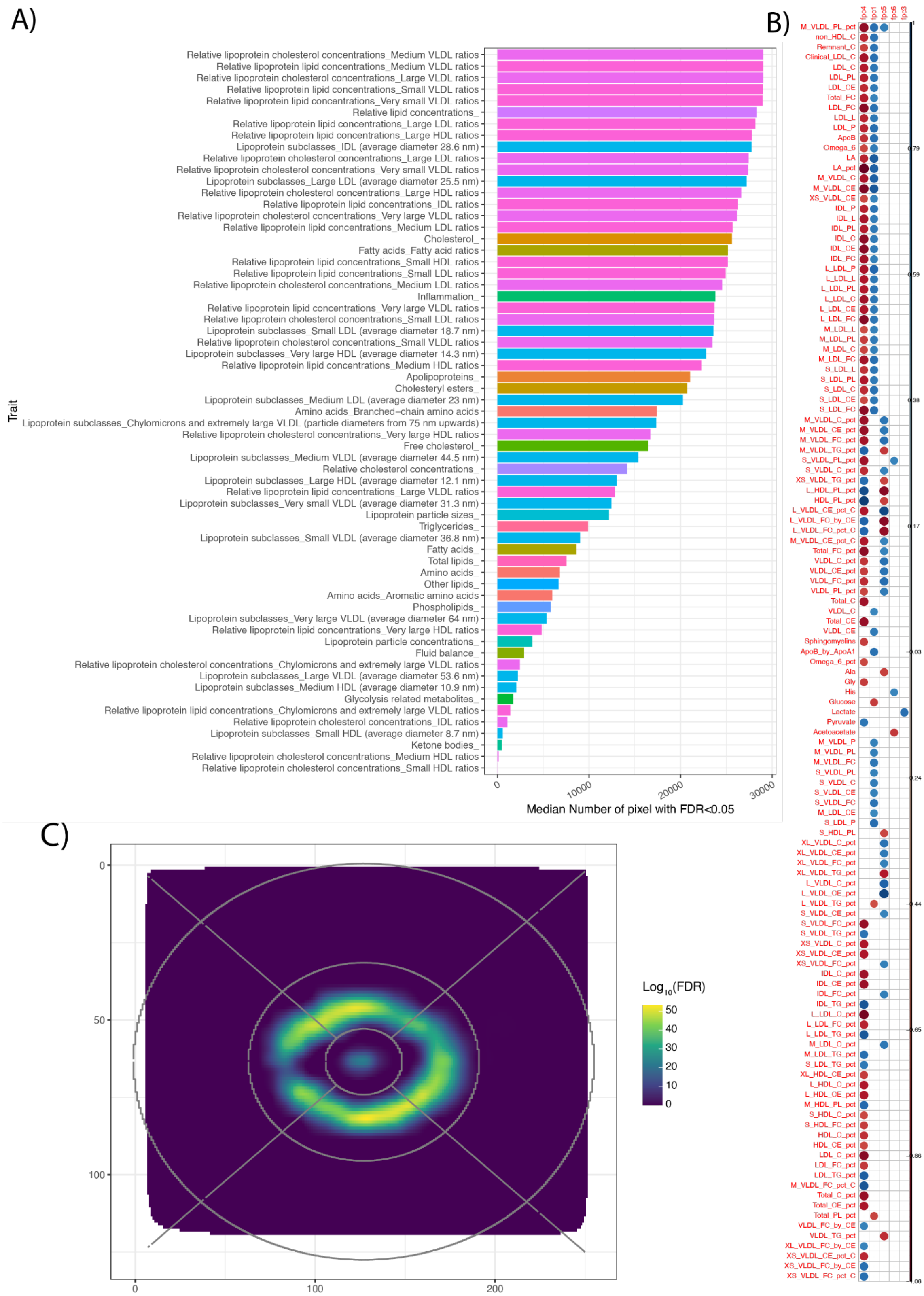


**Extended Data 7. Metabolomics results.** A) Barplot showing the average number of pixels whose thickness was affected by metabolite in each metabolic group. B) 2D smoothed results from ORA analysis on age-interaction metabolic results. C) Heatmap showing significant association between metabolic measures and thickness fPCs. Full metabolites names available in Supplementary Table 14.

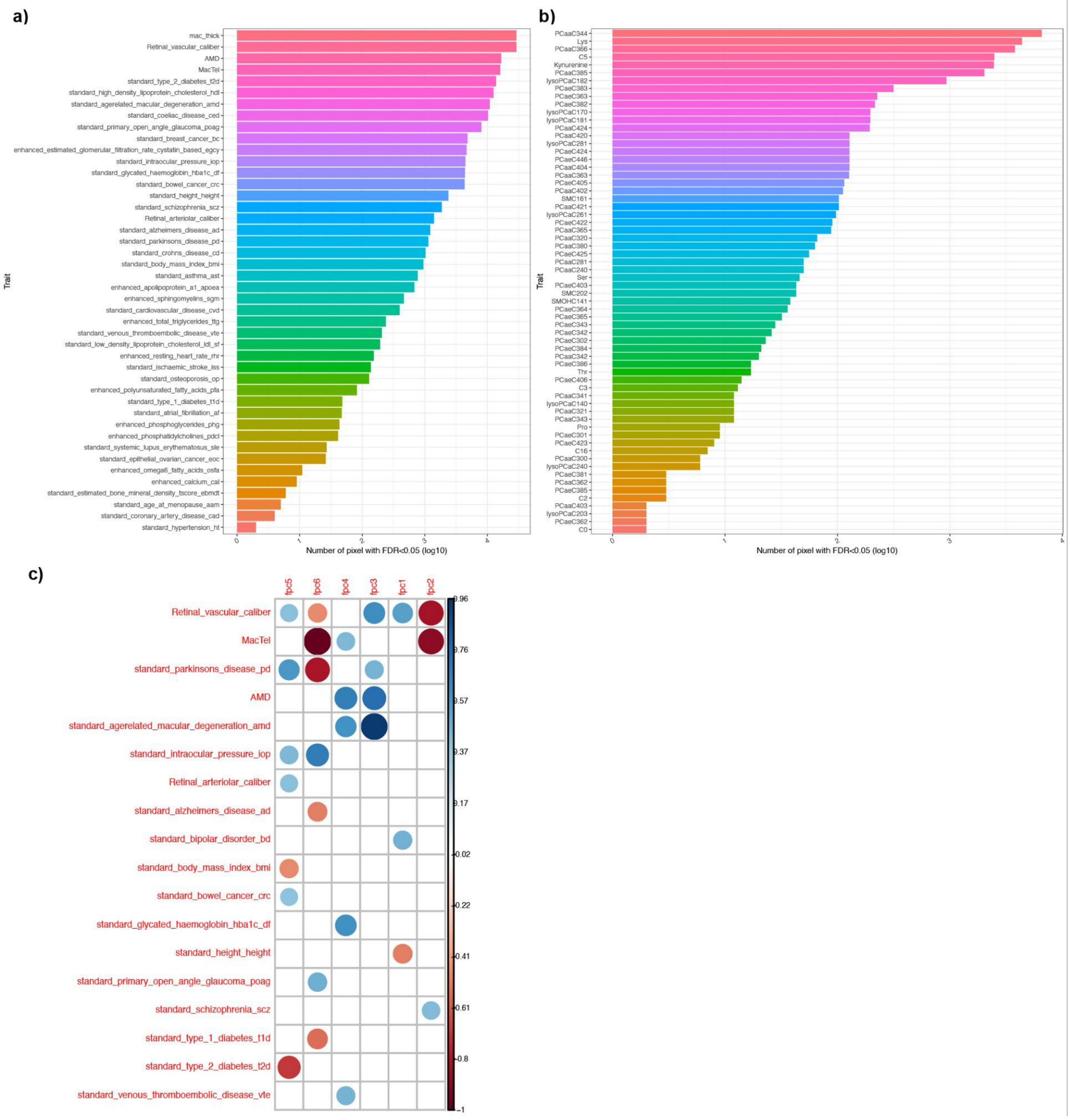


**Extended Data 8. Polygenic risk scores results.** A) Barplot showing the number of pixels which thickness was affected by each significant trait polygenic risk score. B) Barplot showing the number of pixels which thickness was affected by each significant metabolic polygenic risk score (full metabolites names available in Supplementary Table 15). C) Heatmap showing significant association between trait polygenic risk scores and thickness fPCs (average retinal thickness PRS removed given extreme effect on all fPCs).

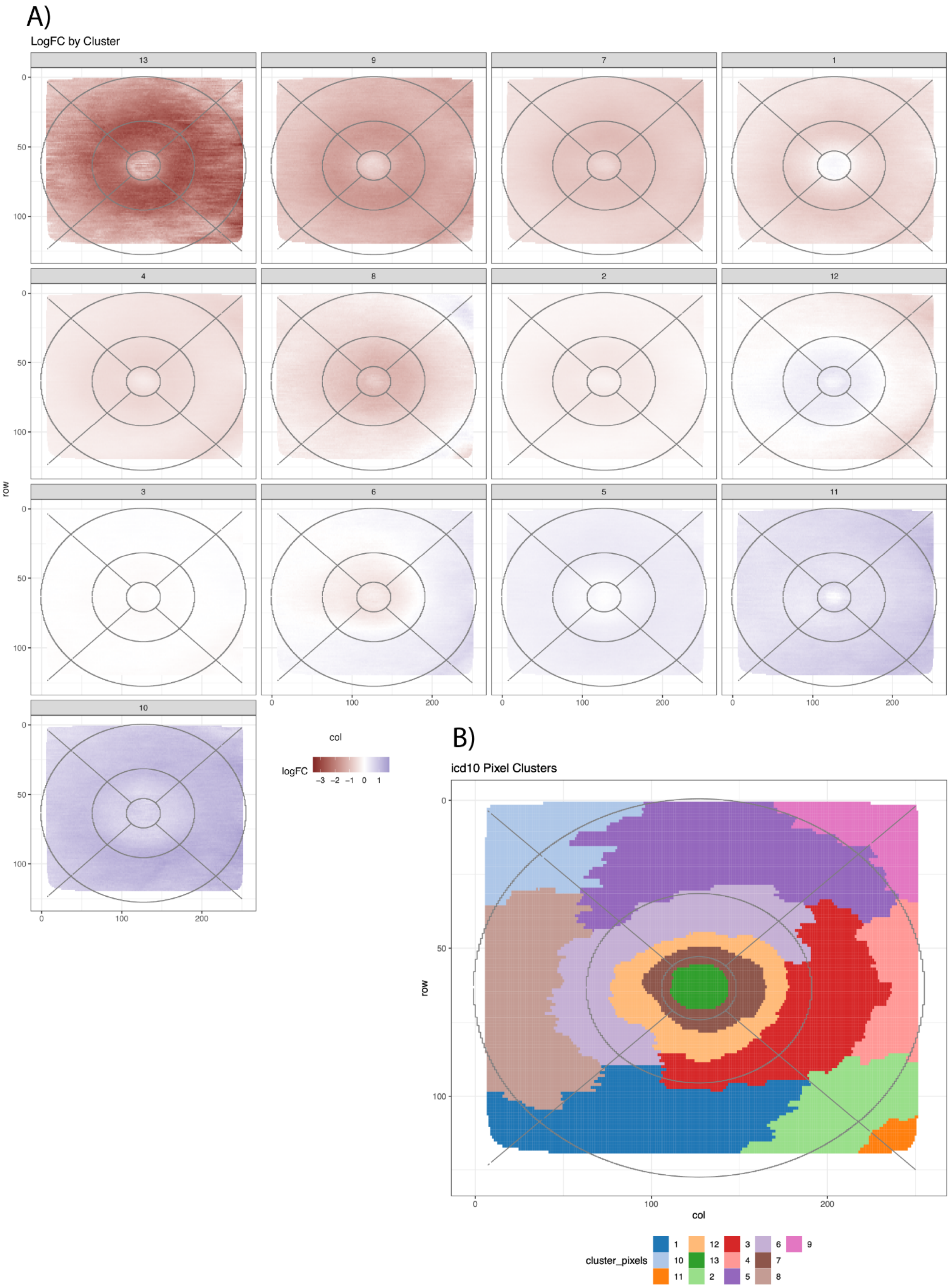


**Extended Data 9. Clustering based on ICD10 results.** Cluster memberships can be found in Supplementary Table 17. A) Pixel-wise average effect across ICD10 within each detected cluster. B) Pixel clusters based on ICD10 diagnoses effects.

A)

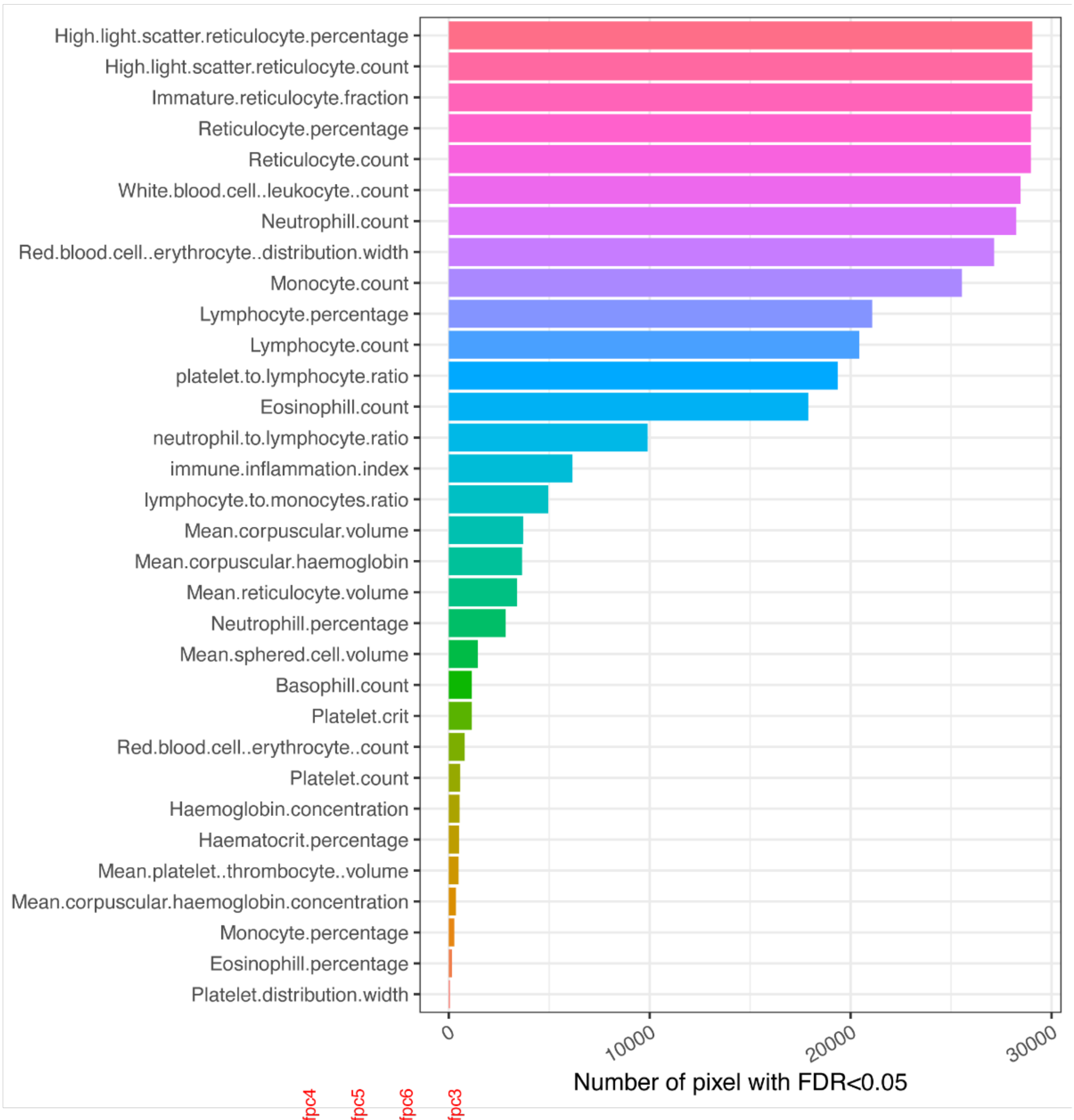

High.light.scatter.reticulocyte.percentage
High.light.scatter.reticulocyte.count
Immature.reticulocyte.fraction
Reticulocyte.percentage
Reticulocyte.count
White.blood.cell..leukocyte..count
Neutrophill.count
Red.blood.cell..erythrocyte..distribution.width
Monocyte.count
Lymphocyte.percentage
Lymphocyte.count
platelet.to.lymphocyte.ratio
Eosinophill.count
neutrophil.to.lymphocyte.ratio
immune.inflammation.index
lymphocyte.to.monocytes.ratio
Mean.corpuscular.volume
Mean.corpuscular.haemoglobin
Mean.reticulocyte.volume
Neutrophill.percentage
Mean.sphered.cell.volume
Basophill.count
Platelet.crit
Red.blood.cell..erythrocyte..count
Platelet.count
Haemoglobin.concentration
Haematocrit.percentage
Mean.platelet..thrombocyte..volume
Mean.corpuscular.haemoglobin.concentration
Monocyte.percentage
Eosinophill.percentage
Platelet.distribution.width
0
10000
20000
30000
Number of pixel with FDR<0.05


B)

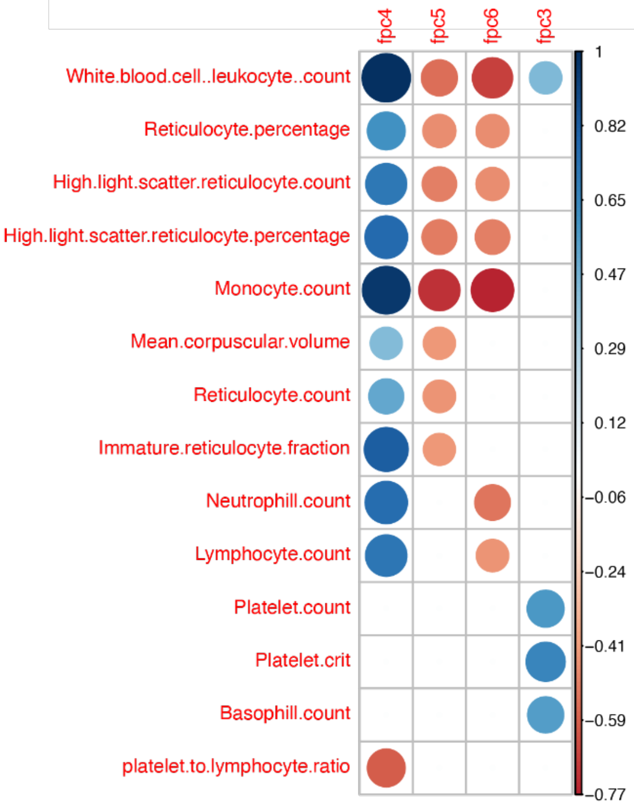

fpc4
fpc5
fpc6
fpc3
White.blood.cell..leukocyte..count
Reticulocyte.percentage
High.light.scatter.reticulocyte.count
High.light.scatter.reticulocyte.percentage
Monocyte.count
Mean.corpuscular.volume
Reticulocyte.count
Immature.reticulocyte.fraction
Neutrophill.count
Lymphocyte.count
Platelet.count
Platelet.crit
Basophill.count
platelet.to.lymphocyte.ratio
1
0.82
0.65
0.47
0.29
0.12
−0.06
−0.24
−0.41
−0.59
−0.77

**Extended Data 10. Blood and inflammation markers results.** A) Barplot showing the number of pixels which thickness was affected by each significant blood or inflammation marker. B) Heatmap showing significant association between significant blood / inflammation marker and thickness fPCs.

### Supplementary Figures

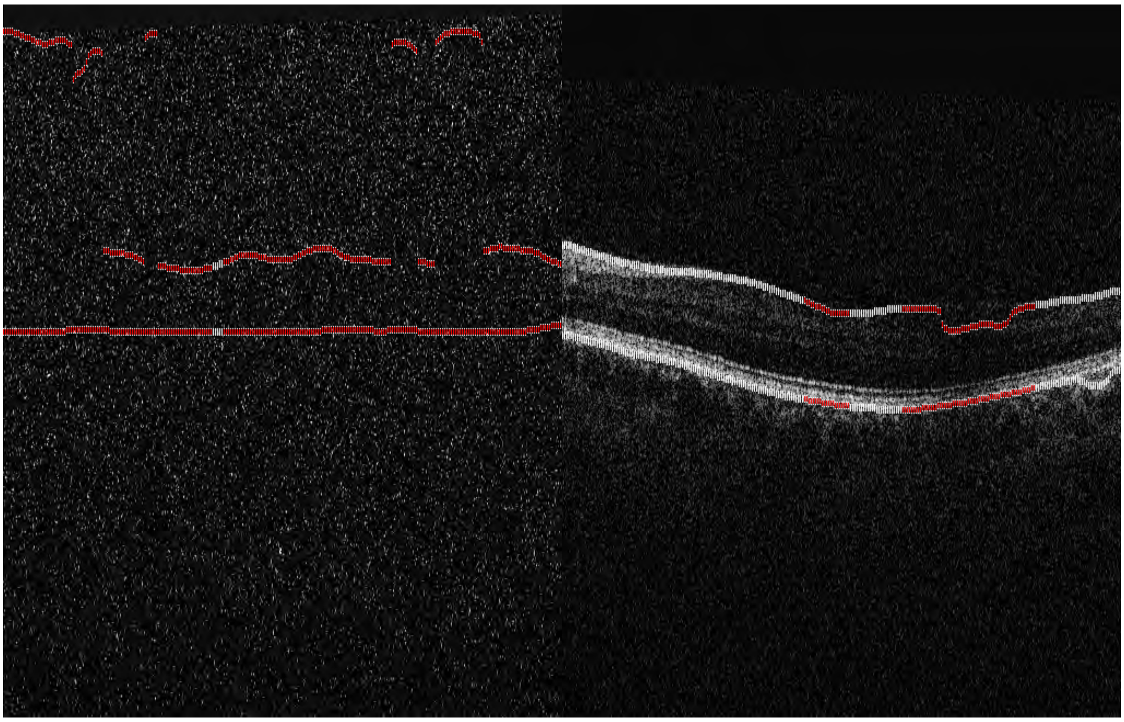

**Supplementary Figure 1.** Quality exclusion criterion 1 - faint B-scans. **A.** Too faint often happens when no retina is imaged, **B.** Some parts of the retina are too faint.

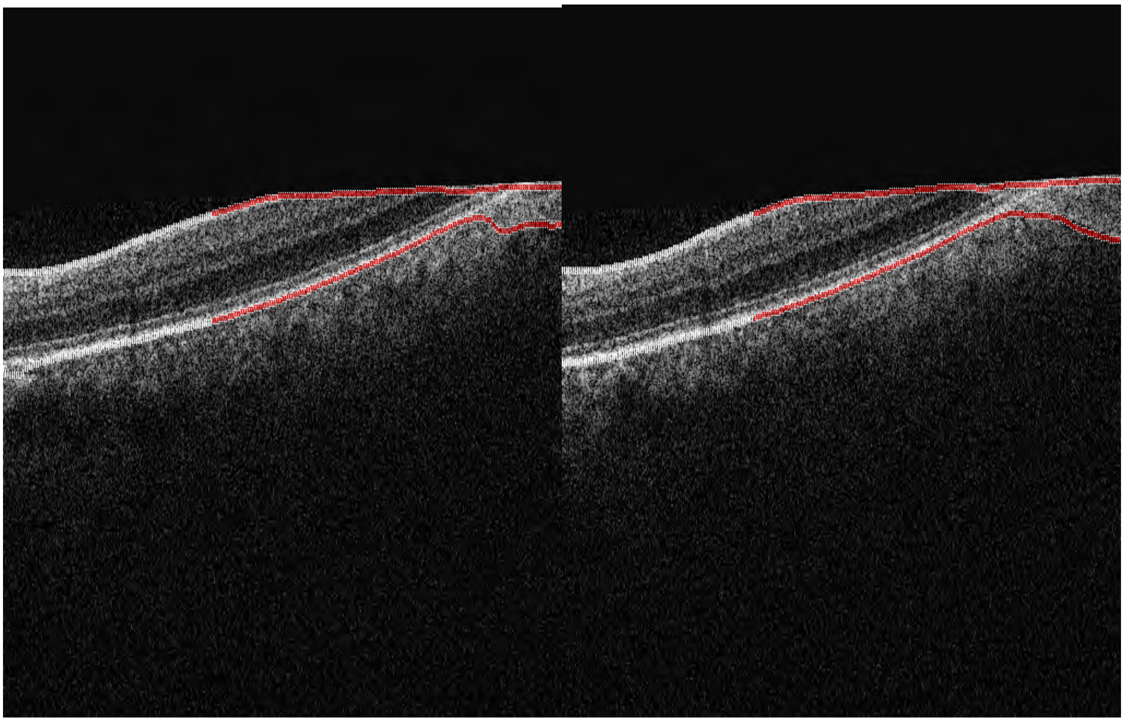

**Supplementary Figure 2.** Quality exclusion criterion 2 - Thickness measurements too thin in scan region. **A.** and **B.** The retina is too thin when it is cut off.

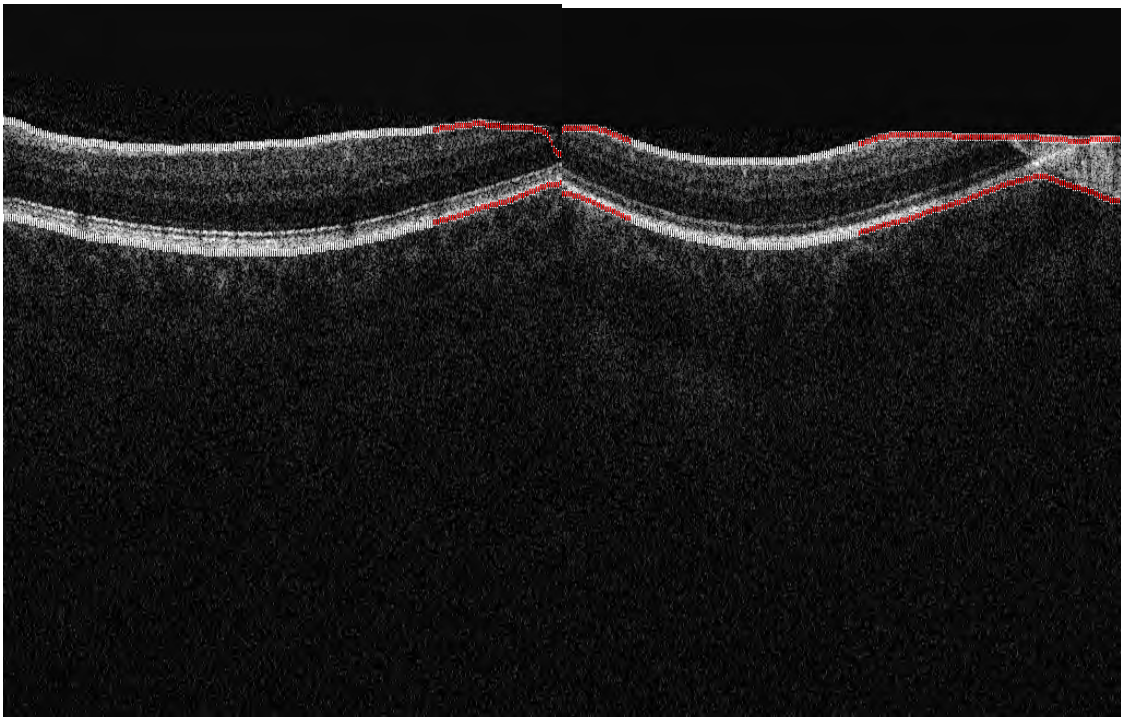

**Supplementary Figure 3.** Quality exclusion criterion 3 - thickness too thin at specific locations. **a.** and **b.** When the retina is cut off, there is no background reflectivity near the ILM or RPE layer boundaries.

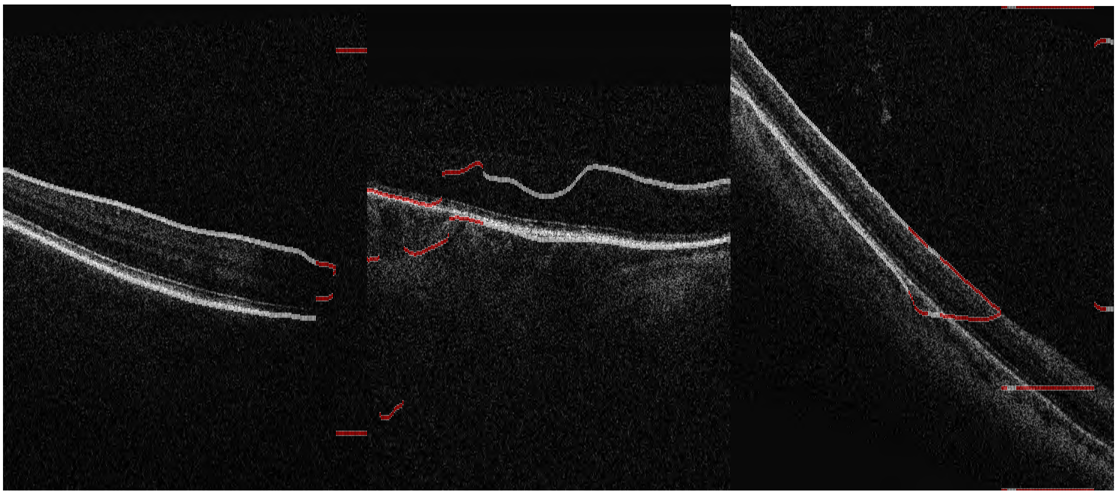

**Supplementary Figure 4.** Quality exclusion criterion 4 - Discontinuities in ILM-RPE segmentation. **Figures 4a** and **4b** are due to OCT quality and **4c** due to algorithm failures.

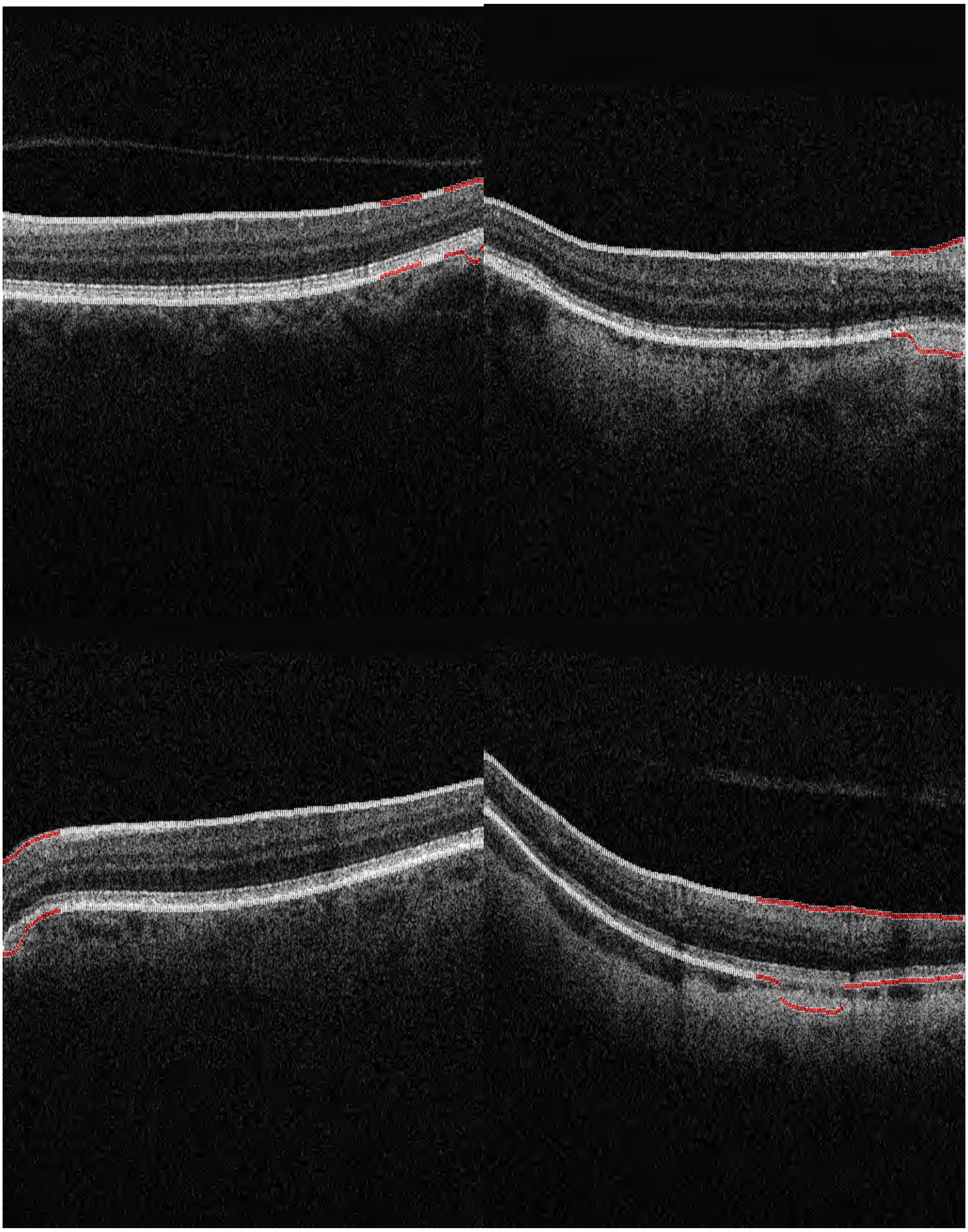

**Supplementary Figure 5.** Quality exclusion criterion 5 - Location thickness discontinuities. **A-D** show examples of anomalies detected by local variability in the ILM-RPE segmentation.

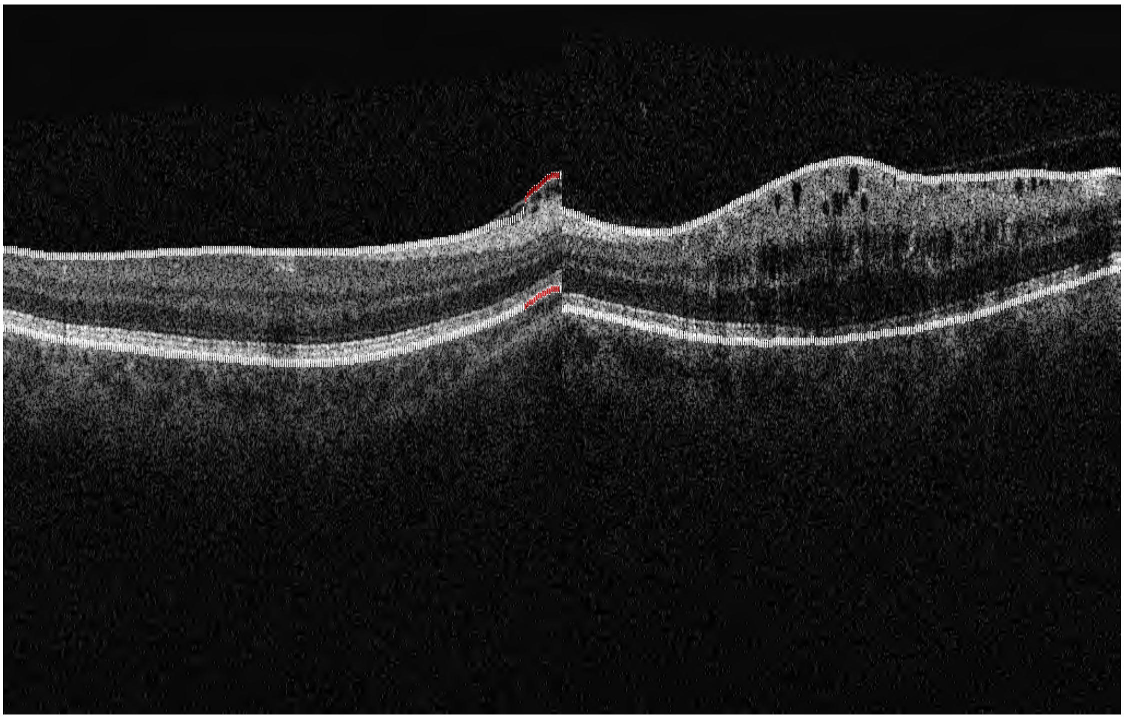

**Supplementary Figure 6.** Quality exclusion criterion 6 - undocumented retinal disease. A and B show examples of diseased retinas flagged by the quality control step. A was flagged by large local deviations, and **B** by too thick a retina due to gaps in the retinal layers.

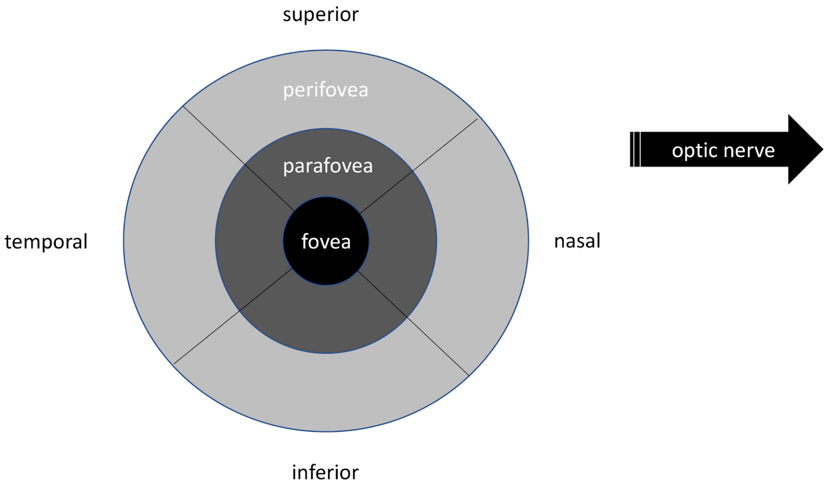


**Supplementary Figure 7.** Early treatment of diabetic retinopathy (ETDRS) grid with orientation used for all retinal images.

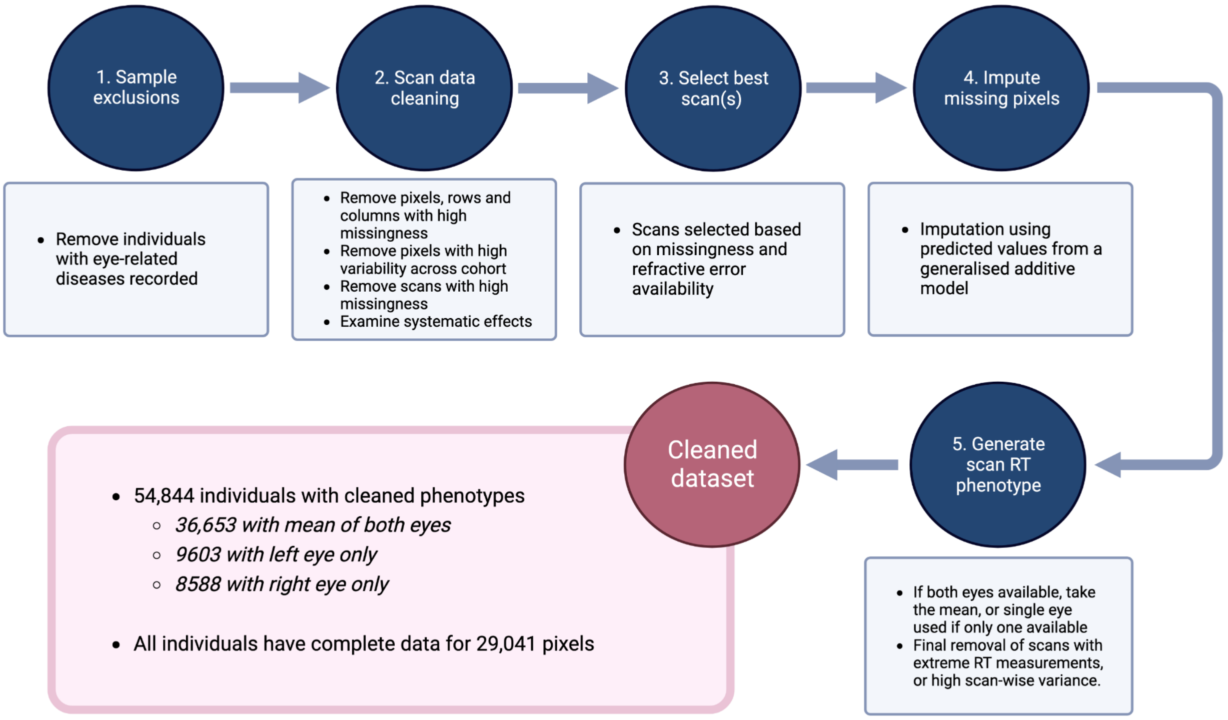


**Supplementary Figure 8.** Quality control filtering. Depiction of quality control filtering at both individual and pixel levels.

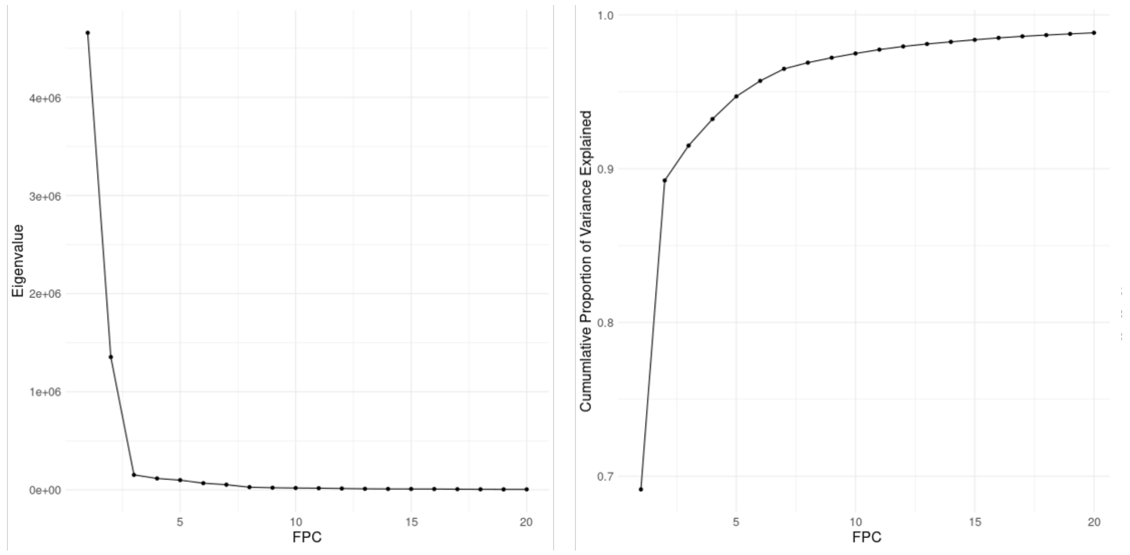


**Supplementary Figure 9.** Two dimensional functional principal component analysis. Scree plot (A), and plot of cumulative variance explained (B) for RT data for 2D fPC dimensionality reduction approach. Six 2D-fPCs capture 95% of all of the RT variation.

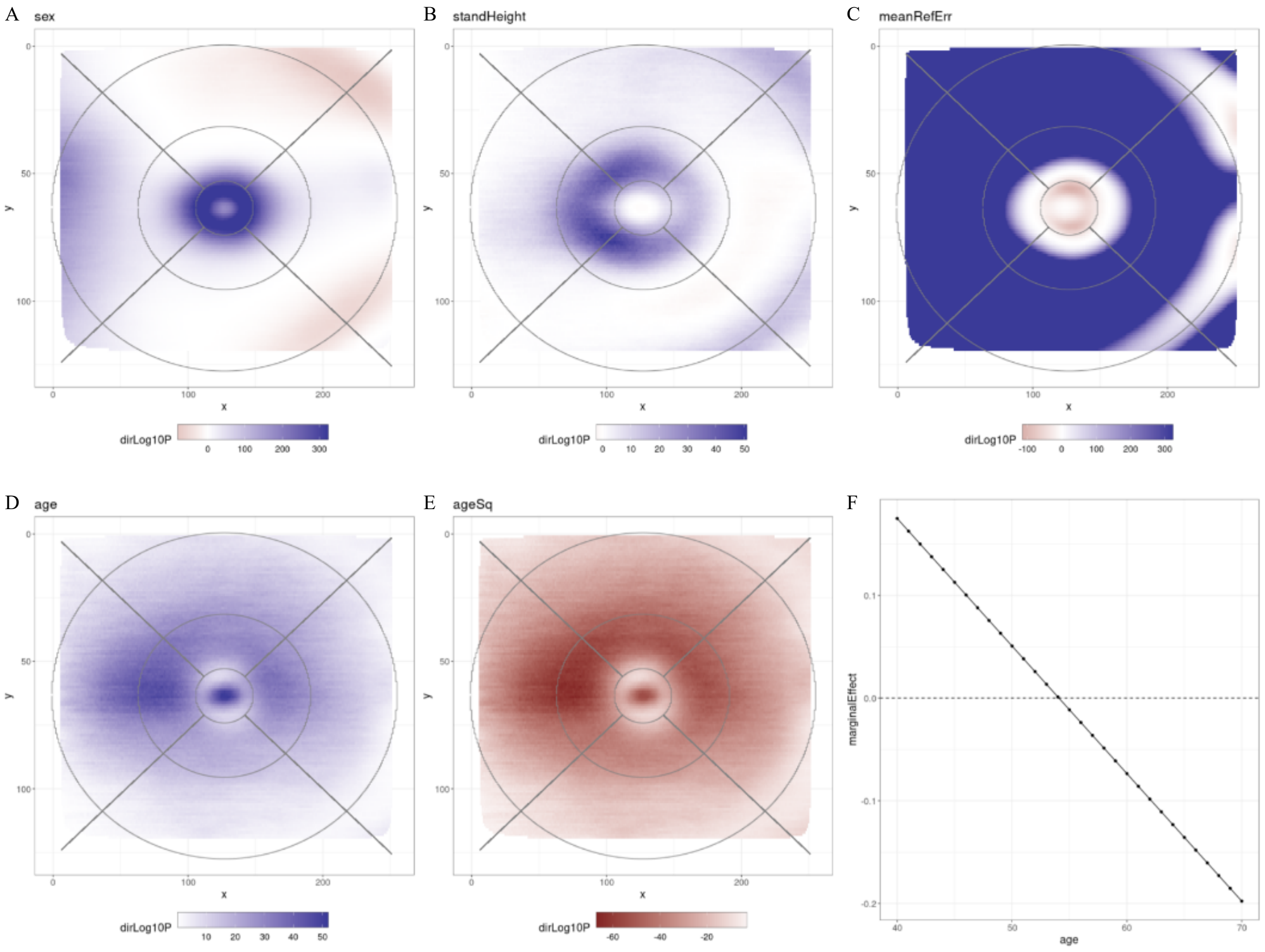


**Supplementary Figure 10:** Pixel-level associations with basic characteristics. Pixel-level associations with A) sex B) standing Height C) refractive error (spherical equivalent), D) age and E) age-squared. F) shows the marginal effect of age for central foveal pixel (64_128).

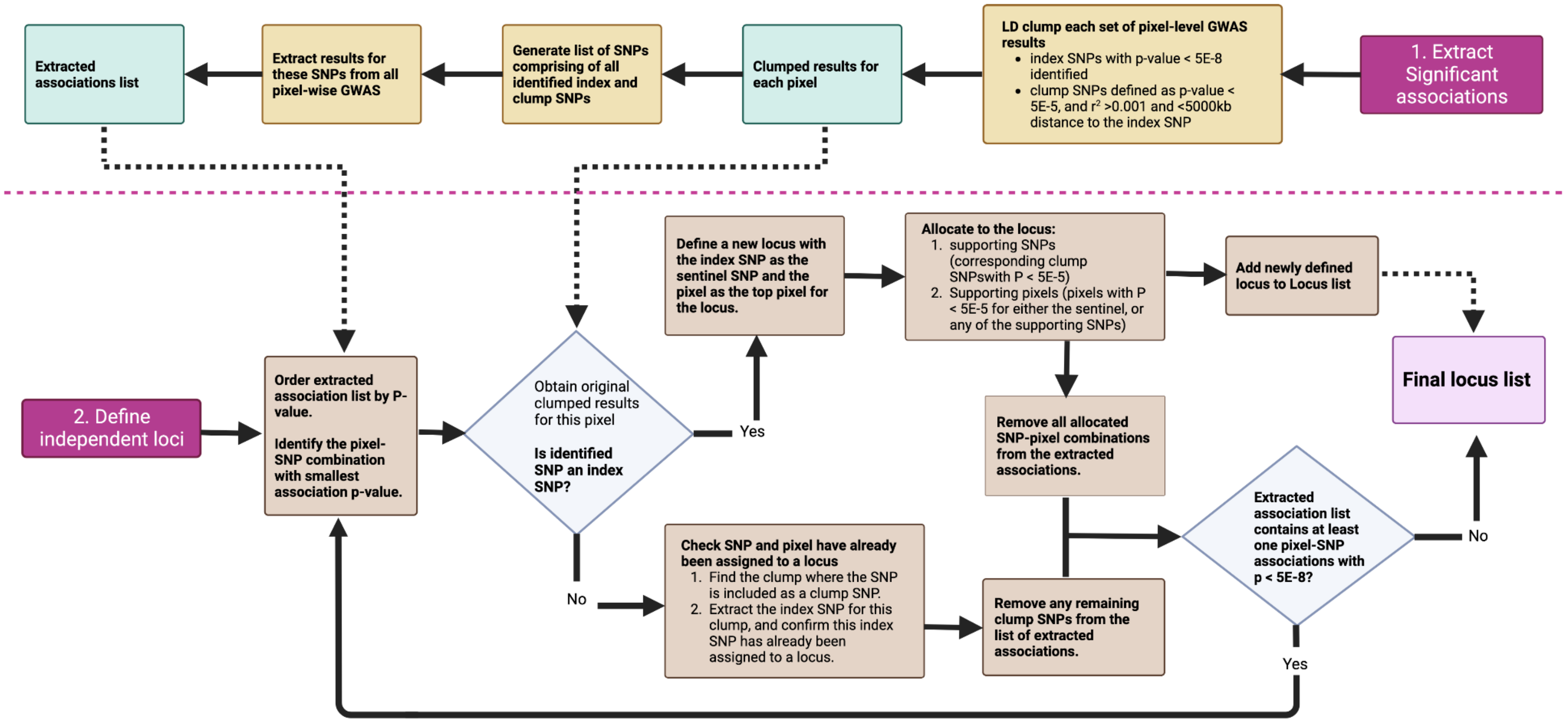


**Supplementary Figure 11.** Procedure for collation of pixel-level GWAS results. Flowchart of iterative procedure developed to summarise GWAS results from all 29,041 RT pixels

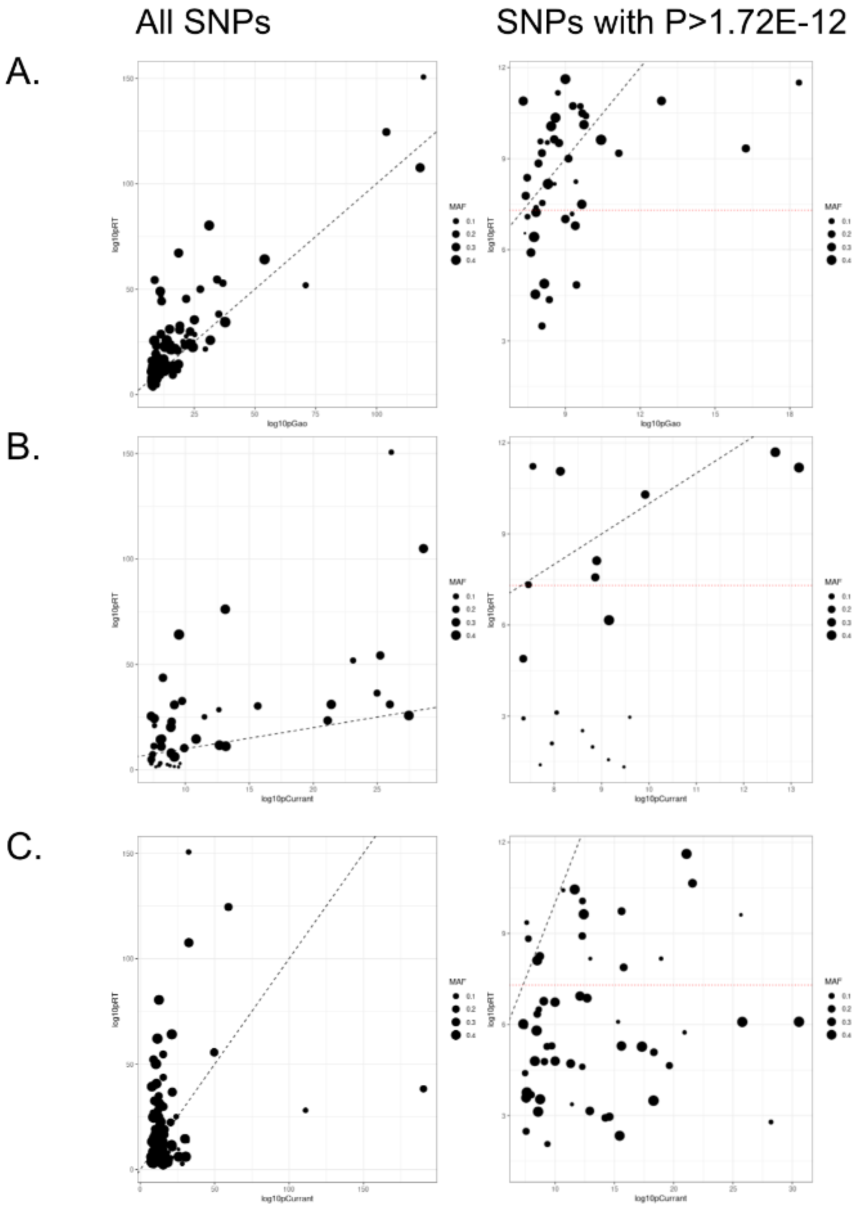


**Supplementary Figure 12.** Comparison with previously identified GWAS loci. Shown are comparison of effect sizes for SNPs identified in previous studies (A. (Gao et al. 2018); B. (Currant et al. 2021) C. (Currant et al. 2023)), vs the smallest p-value from the pixel-level or fPC analyses. Data points are sized based on SNP MAF. The left pot shows all SNPs. The right plot shows SNPs not meeting the pixel-level Bonferroni adjusted significance level. The red dotted line shows the standard genome-wide significance threshold (P<5E-8).

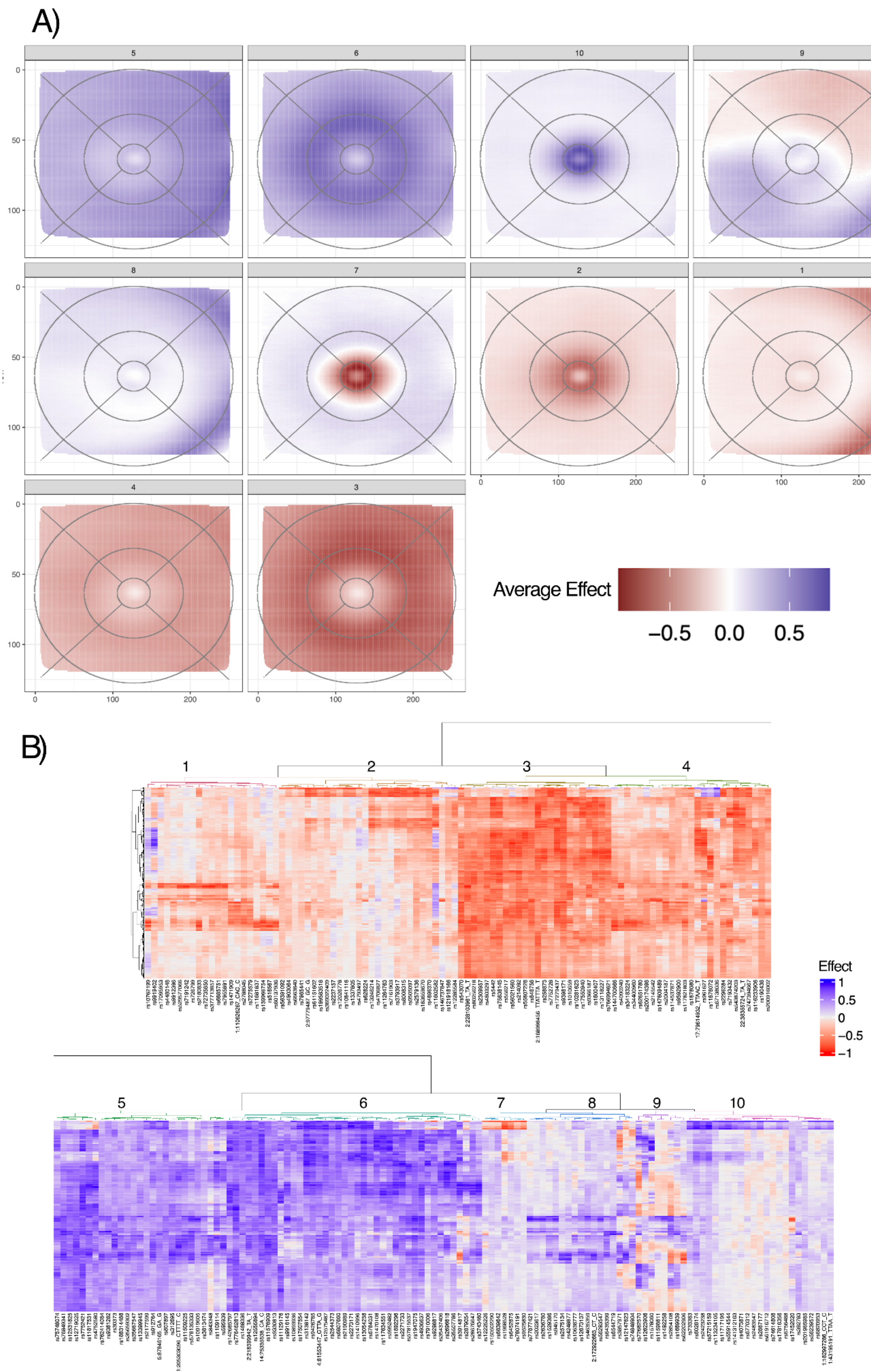


**Supplementary Figure 13.** Clustering of SNPs identified through the pixel-level analyses, based on effects across all 29,041 pixels. A. Pixel-wise average effect across SNPs within each detected cluster. B) Heatmap showing SNPs clusters and their effect on a subset of pixels.

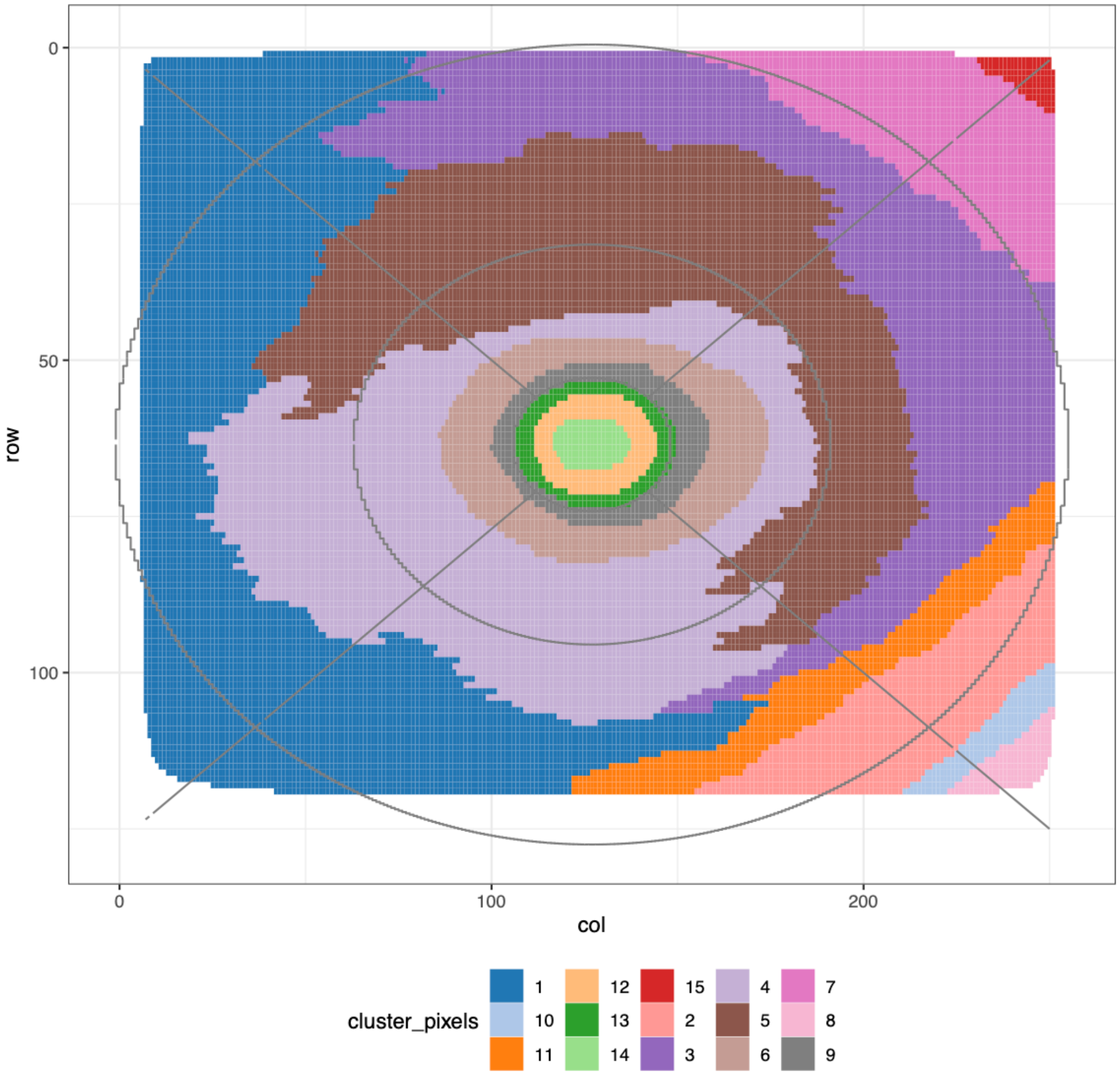


**Supplementary Figure 14.** Clustering of pixels, based on SNP effects for loci identified through the pixel-level analyses.

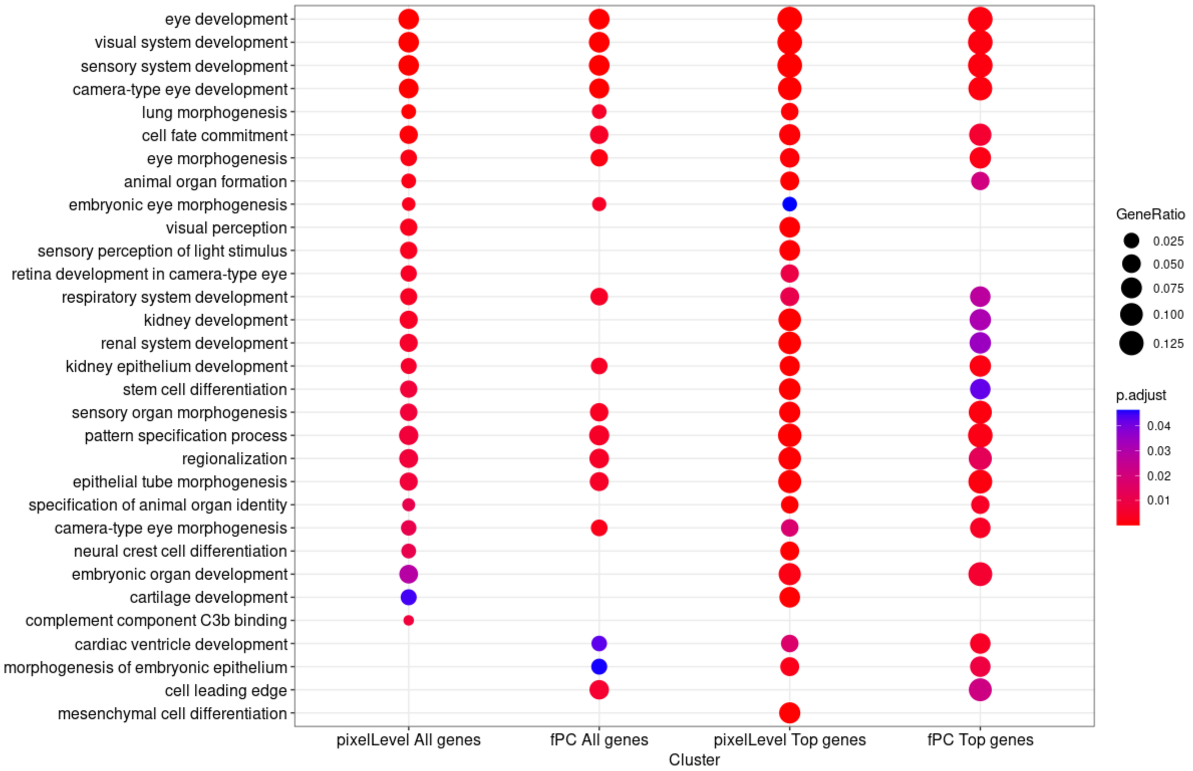


**Supp Figure 14.** Gene Ontology Over-Representation Analysis

Shown are gene Ontology, over-enrichment analysis results for four gene sets: all candidate genes implicated via the pixel-level and fPC locus to gene mapping, and the “Top genes” for each analysis, defined as the gene(s) with the most lines of evidence for each locus (i.e. highest “evidenceScore”, Supplementary Table 10).
